# A Zero-Burden Sleep Foundation Model Built on Cardiorespiratory Signals from 1,400,000+ Hours of Multi-Ethnic Sleep Recordings

**DOI:** 10.1101/2025.09.06.25335216

**Authors:** Guangkun Nie, Xuesong Chen, Yichen Wang, Jingxu Chen, Yunhan Shi, Jianwen Zhong, Xiaocheng Fang, Jie Shi, Chun-feng Liu, Bei Huang, Yaping Liu, Jihui Zhang, Yi Fang, Jiong Chen, Robert J. Thomas, Weijun Huang, Zengrui Jin, Fei Lei, Leilei Wang, Rui Zhao, Chao Zhang, Kaibing Chen, Dongsheng Lv, Wei Chen, Hongliang Yi, Jun Liu, Yun-Kwok Wing, Hongyan Li, M. Brandon Westover, Lin Lu, Xiangdong Tang, Shankai Yin, Yanru Li, Shenda Hong, Yue Leng, the AISleep 365 Consortium

## Abstract

**BACKGROUND:** Sleep disorders pose a major global health burden and are associated with a wide range of adverse health outcomes. Polysomnography (PSG) is the gold standard for sleep assessment, but it remains impractical for home-based or long-term monitoring. We investigated whether overnight cardiorespiratory signals, which can be acquired with zero-burden to users, can enable accurate sleep assessment at scale and capture broader dimensions of multi-organ health when leveraged through foundation model approaches.

**METHODS:** We present SleepFounder, a foundation model for zero-burden sleep monitoring built upon cardiorespiratory signals. SleepFounder was developed on the largest curated multi-ethnic sleep dataset to date, comprising over 1.4 million hours of recordings from 34 cohorts across the United States and China. We evaluated SleepFounder across diverse downstream tasks, including conventional sleep analysis, demographic profiling, and multi-organ disease detection and prediction. We further conducted a real-world external validation study using multi-center ballistocardiography (BCG) data collected with a custom-developed under-pillow BCG monitoring system.

**RESULTS:** SleepFounder achieved strong performance across downstream tasks, outperforming baseline models in 19 of 21 dataset-task pairs spanning conventional sleep analysis and demographic profiling. For example, when averaged across three to five external datasets, SleepFounder achieved a Cohen’s kappa of 0.651 [0.649-0.652] for five-class sleep staging and an AUROC of 0.912 [0.909-0.915] for moderate-to-severe obstructive sleep apnea (OSA) detection. Beyond sleep-specific tasks, SleepFounder demonstrated broad capability for multi-organ disease detection and future disease prediction. Averaged across three external cohorts, it achieved AUROCs > 0.75 for 172 disease phenotypes and concordance indices (C-indices) > 0.75 for 100 phenotypes spanning 16 phecode chapters, including clinically important conditions such as dementia (AUROC=0.850 [0.836-0.862], C-index=0.845 [0.833-0.855]), Parkinson’s disease (AUROC=0.846 [0.832-0.860], C-index=0.823 [0.803-0.842]), congestive heart failure (AUROC=0.840 [0.834-0.846], C-index=0.768 [0.759-0.778]), atrial fibrillation (AUROC=0.835 [0.828-0.842], C-index=0.762 [0.751-0.773]), and respiratory failure (AUROC=0.822 [0.810-0.833], C-index=0.778 [0.767-0.789]). Importantly, SleepFounder generalized effectively to real-world BCG deployment, achieving a Cohen’s kappa of 0.621 [0.614-0.627] for five-class sleep staging, an AUROC of 0.949 [0.940-0.958] for moderate-to-severe OSA detection, an age-prediction MAE of 6.533 [6.278-6.792] years, and a sex-classification AUROC of 0.852 [0.834-0.869] in external multi-center validation.

**CONCLUSIONS:** SleepFounder establishes a foundation model that learns from cardiorespiratory signals to enable accurate and scalable sleep assessment. By linking sleep physiology with multi-organ health, it demonstrates the potential to bridge clinical and home settings, while suggesting that signals traditionally used for sleep monitoring may also serve as powerful biomarkers of systemic function and disease risk. These findings highlight a new paradigm for home-based sleep and health monitoring in real-world settings.

## Introduction

Sleep is essential for overall health, with poor sleep quality linked to increased risks of cardiovascular diseases (CVDs)^1,2,3,4^, metabolic dysfunction^5,6^, neurological disorders^7,8^, and psychiatric conditions^9,10^. Sleep disorders, such as obstructive sleep apnea (OSA), are a growing global health concern. For example, an estimated 936 million adults aged 30-69 worldwide suffer from mild to severe sleep apnea^11^. In the United States, 80% of OSA cases among nearly 85 million affected individuals remain undiagnosed^12,13^. Polysomnography (PSG), the gold standard for sleep analysis, captures high-fidelity physiological signals such as electroencephalography (EEG), respiration, electrocardiography (ECG), and blood oxygen saturation (SpO_2_). Despite its accuracy, PSG is resource-intensive and disrupts natural sleep, making it unsuitable for home-based or long-term monitoring^14,15^. There is an urgent need for cost-effective solutions that enable reliable sleep monitoring without imposing additional burden on users.

Recent studies^16,17,18^ have explored the use of reduced subsets of PSG channels to enable lower-burden sleep assessment. In particular, deep learning methods applied to cardiorespiratory signals have shown promising performance in key sleep analysis tasks, including sleep staging^19,20,21,22^ and OSA severity/apnea-hypopnea index (AHI) estimation^23,24,25,26^. These signals, which capture the dynamics of the cardiovascular and respiratory systems, are strongly linked to sleep patterns and overall physiological states. Importantly, they represent a shared physiological modality captured in both clinical PSG and home-based systems such as ballistocardiography (BCG)^22,26^, creating the opportunity to assemble large, heterogeneous datasets well suited for advanced deep learning techniques. However, most prior studies have overlooked this shared characteristic, relying on limited dataset size and demographic diversity, thereby constraining the accuracy, generalizability, and robustness of existing approaches. Foundation models, which leverage large-scale pretraining and generalize across diverse downstream tasks, have shown considerable promise in the medical domain, including computational pathology^27,28^, echocardiography^29^, and physiological signal analysis^30,31,32,33,**Error!**^ **^Reference^ ^source^ ^not^ ^found.^**. Building on these advances, we propose that large-scale, physiologically salient cardiorespiratory data—derived from both PSG and home-based devices—combined with foundation model techniques, can enable accurate, scalable sleep assessment and extend toward broader multi-organ health evaluation.

In this study, we introduce SleepFounder, a foundation model for accurate, zero-burden sleep monitoring based solely on cardiorespiratory signals (**Figure 1**). The model was developed on the largest curated multi-ethnic sleep dataset to date, totaling over 1.4 million hours of sleep recordings from 34 cohorts across the United States and China. During pretraining, SleepFounder was guided by two physiology-inspired pretext tasks, EEG spectrogram reconstruction and oxygen desaturation prediction, which enabled the model to extract physiologically meaningful information embedded in cardiorespiratory signals and to internalize latent mechanisms linking these dynamics to sleep. While prior foundation models such as SleepFM**^Error!^ ^Reference^ ^source^ ^not^ ^found.^** have advanced sleep representation learning using multi-channel PSG data, SleepFounder focuses on widely accessible cardiorespiratory signals, enabling accurate and flexible sleep analysis with potential for population-scale deployment.

**Figure 1.**
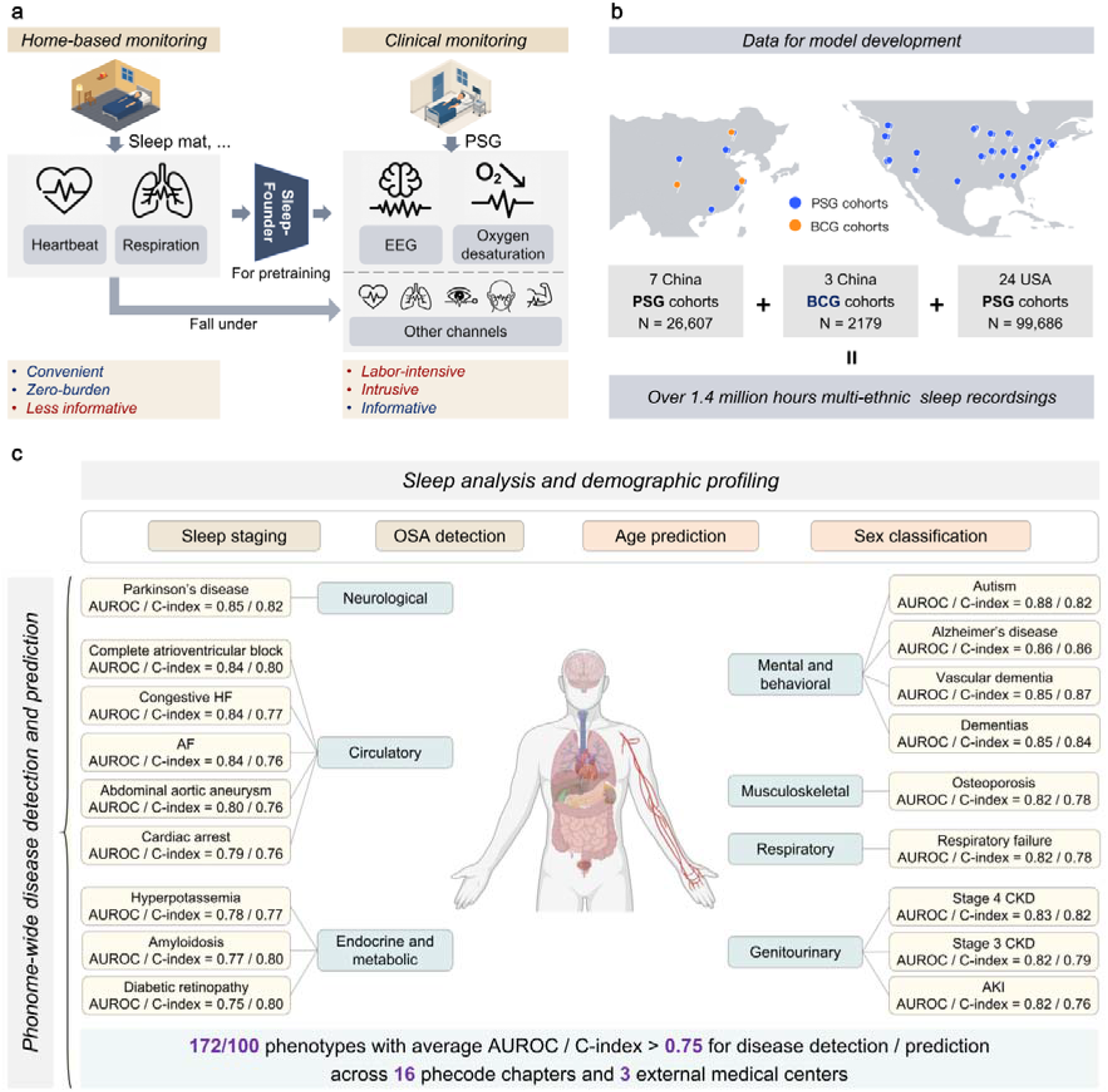
Overview of the proposed SleepFounder framework. (a) SleepFounder leverages respiratory and heartbeat signals shared between clinical PSG systems and contactless home-based sleep monitoring devices. During pretraining, the model learns to reconstruct EEG and oxygen desaturation signals, using informative modalities to guide the representation learning of less informative ones. (b) SleepFounder was developed using large-scale, multi-ethnic sleep datasets from China and the United States, comprising seven Chinese PSG cohorts, three real-world BCG cohorts, and twenty-four USA PSG cohorts, totaling over 1.4 million hours of recorded sleep data. (c) SleepFounder was evaluated across multiple downstream tasks, including conventional sleep analysis and demographic profiling. Additionally, in phenome-wide analyses of multi-organ disease detection and future disease prediction, SleepFounder achieved strong performance across 16 phecode chapters, with averaged AUROCs > 0.75 for 172 prevalent disease phenotypes and averaged C-indices > 0.75 for 100 incident disease phenotypes across three external medical centers. Representative phenotypes are shown. PSG, polysomnography; EEG, electroencephalography; BCG, ballistocardiography; OSA, obstructive sleep apnea; AUROC, area under the receiver operating characteristic curve; C-index, concordance index; HF, heart failure; AF, atrial fibrillation; CKD, chronic kidney disease; AKI, acute kidney injury.

## Methods

### Study design and oversight

This study used both public and private datasets and was conducted in accordance with the principles of the Declaration of Helsinki. Ethical approval for the overall project was obtained from the Biomedical Ethics Review Committee of Peking University (IRB00001052-23207). In addition, local institutional review board approvals were obtained from all participating clinical centers, including the Affiliated Hospital of Gansu University of Chinese Medicine (AHGUCM), Beijing Huilongguan Hospital (BHH), Inner Mongolia Mental Health Center (IMMHC), Shenzhen Hospital of Southern Medical University (SHSMU), Beijing Tongren Hospital (BTH affiliated to Capital Medical University), Sir Run Run Shaw Hospital (SRRSH affiliated to Zhejiang University School of Medicine), Shanghai Sixth People’s Hospital (SSPH affiliated to Shanghai Jiao Tong University School of Medicine), and the Sleep Medicine Center of West China Hospital (WCH affiliated to Sichuan University). As only de-identified retrospective data were used, the requirement for informed consent was waived by each institutional review board.

### Dataset allocation and evaluation framework

A total of 34 cohorts were used for SleepFounder development, including 26 cohorts for the pretraining and downstream fine-tuning stages, five independent PSG cohorts for external evaluation, and three BCG cohorts for real-world validation. Dataset inclusion followed predefined modality-specific criteria. For PSG data, recordings were included if both ECG and either chest or abdominal respiratory signals were available, with detectable ECG R peaks required for RR interval extraction. For BCG data, inclusion was determined using the signal quality score proposed in SleepNetZero^34^, with only recordings scoring above 0.5 retained for analysis. An overview of dataset allocation and the overall development workflow is provided in **Figure 2**, and detailed cohort descriptions are available in Supplementary Methods, Section 1.

**Figure 2.**
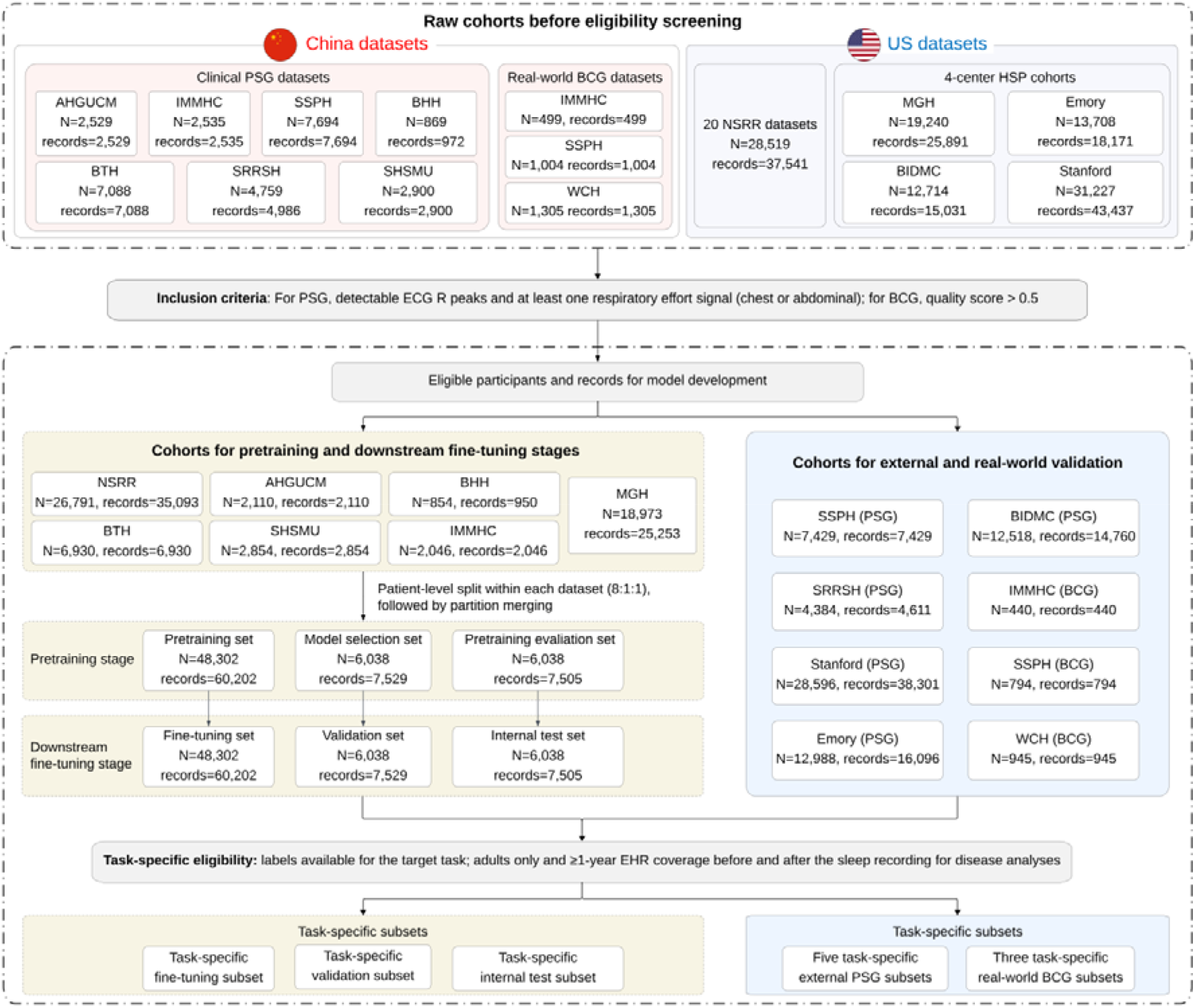
Data flowchart of SleepFounder development. Multi-center PSG and BCG cohorts from the United States and China were screened according to modality-specific inclusion criteria. Eligible participants and recordings were assigned to cohorts for the pretraining and downstream fine-tuning stages or to independent cohorts for external and real-world validation. Participant-level splits were created for the pretraining stage and subsequently reused in the downstream fine-tuning stage as the fine-tuning, validation, and internal test sets. For both downstream fine-tuning and independent evaluation in external PSG and real-world BCG cohorts, task-specific subsets were further derived from the corresponding datasets according to predefined eligibility criteria and label availability. PSG, polysomnography; BCG, ballistocardiography; ECG, electrocardiography; HSP, the Human Sleep Project; AHGUCM, the Affiliated Hospital of Gansu University of Chinese Medicine; IMMHC, Inner Mongolia Mental Health Center; SSPH, Shanghai Sixth People’s Hospital; BHH, Beijing Huilongguan Hospital; BTH, Beijing Tongren Hospital, Capital Medical University; SRRSH, Sir Run Run Shaw Hospital, Zhejiang University School of Medicine; SHSMU, Shenzhen Hospital of Southern Medical University; WCH, the West China Hospital, Sichuan University; NSRR, the National Sleep Research Resource; MGH, Massachusetts General Hospital; BIDMC, Beth Israel Deaconess Medical Center.

The 26 cohorts used for the pretraining and downstream fine-tuning stages comprised 60,378 individuals from the United States and China. The U.S. component consisted of 45,584 participants from 21 cohorts, including the Massachusetts General Hospital (MGH) cohort of the Human Sleep Project (HSP) and 20 datasets from the National Sleep Research Resource (NSRR). The Chinese component comprised 14,794 participants from five clinical cohorts (AHGUCM, BHH, IMMHC, SHSMU, and BTH). For the pretraining stage, these cohorts were randomly partitioned at the participant level into pretraining, model selection, and pretraining evaluation sets in an 8:1:1 ratio, while preserving cohort-specific proportions. The resulting splits were used for gradient-based parameter optimization, checkpoint selection, and internal pretraining evaluation, respectively.

For the downstream fine-tuning stage, the pretraining, model selection, and pretraining evaluation sets were reused as the fine-tuning, validation, and internal test sets, which were used for downstream fine-tuning, checkpoint selection, and internal testing, respectively. Task-specific subsets were then derived from these sets according to predefined eligibility criteria. Specifically, samples were required to have labels available for the corresponding task. For disease detection and future disease prediction, participants were further required to be adults and to have an electronic health record (EHR) observation window extending at least one year before and one year after the sleep recording. Consequently, only the MGH cohort was eligible for model adaptation in disease detection and prediction tasks. Detailed sample sizes for each dataset across downstream tasks are provided in Supplementary Data 3.

External evaluation was conducted using independent cohorts that were entirely excluded from both the pretraining and downstream fine-tuning stages. PSG-based external evaluation included the Beth Israel Deaconess Medical Center (BIDMC), Stanford, and Emory cohorts of HSP, together with two Chinese clinical cohorts (SSPH and SRRSH). Following the same procedure, task-specific subsets were derived from the external cohorts according to label availability. Consequently, the BIDMC, Stanford, and Emory cohorts were used for all downstream tasks, whereas SSPH was additionally used for moderate-to-severe OSA detection and both SSPH and SRRSH were used for age prediction and sex classification (Supplementary Data 3). For disease prediction analyses, the median follow-up durations in the external evaluation cohorts were 3.7, 4.7, and 5.1 years for the BIDMC, Stanford, and Emory cohorts, respectively; the corresponding median follow-up duration in the MGH fine-tuning cohort was 5.4 years. For real-world validation, multi-center BCG datasets were collected concurrently with clinical PSG using our previously developed Five Seasons under-pillow BCG monitoring system^36^ and included cohorts from SSPH, WCH, and IMMHC. Notably, although SSPH contributed to both PSG-based external evaluation and BCG-based real-world validation, the corresponding cohorts consisted of distinct, non-overlapping participants.

### Data preprocessing for pretraining

Modality-specific procedures were applied to extract shared cardiorespiratory components between PSG and BCG recordings, specifically heartbeat and respiratory signals. Overnight EEG and SpO_2_ data were further converted into spectrograms and desaturation curves, respectively, which served as reconstruction targets during pretraining. Detailed preprocessing steps are provided in Supplementary Methods, Section 2.

### Model architecture and pretraining details

SleepFounder consists of a ResNet-based^37^ feature extractor followed by a RoFormer encoder^38^. The model takes overnight heartbeat and respiratory signals as input and adopts an early fusion strategy to facilitate cross-modal feature interaction. During feature extraction, two structurally identical but independently parameterized ResNet branches learn temporal representations from each physiological modality. Each branch begins with a stem module comprising a convolutional layer, batch normalization, ReLU activation, and max-pooling, followed by eight residual blocks that progressively transform raw signals into 512-channel feature representations. Features from the two modalities are fused through element-wise summation and passed to a RoFormer encoder to capture long-range temporal dependencies. The RoFormer comprises six stacked layers, each with eight attention heads, a hidden dimension of 512, and a feedforward dimension of 2048. The overall architecture and detailed configurations are provided in Supplementary Methods, Section 5.

The pretraining stage is designed to enable the model to learn semantically meaningful and physiologically grounded representations by jointly optimizing two pretext tasks: EEG spectrogram reconstruction and oxygen desaturation prediction. These tasks capture complementary aspects of sleep physiology, with the former focusing on neural dynamics and the latter on cardiorespiratory patterns. Features extracted by the RoFormer encoder are passed through two task-specific linear heads. The EEG reconstruction head is trained using mean squared error between predicted and ground-truth spectrograms. The oxygen desaturation head predicts continuous SpO_2_ trajectories over 30-second intervals, with a loss combining Wasserstein distance, which captures cumulative deviations over time, and Pearson correlation loss, which promotes linear alignment with ground-truth trends. The combined SpO_2_ loss is defined as *L*_SpO2_ = *λ*_Pearson_*L*_Pearson_ + *λ*_Wasserstein_*L*_Wasserstein_, where λ_Pearson_ and λ_Wasserstein_ were set to 0.1 and 0.9, respectively. The overall pretraining objective integrates both tasks as a weighted sum, *L*_total_ = *L*_EEG_ + *λ*_SpO2_ *L*_SpO2_, with the weight *λ*_SpO2_ set to 0.1. Additional implementation details are provided in Supplementary Methods, Section 3.

During pretraining, the model was trained on four NVIDIA H20 GPUs using the Adam optimizer, with a batch size of 65 and an initial learning rate of 1e-4. Training was conducted for up to 20 epochs with early stopping based on validation loss, using a patience of 10 epochs.

### Fine-tuning strategy

To adapt SleepFounder for downstream tasks, the pretraining heads for EEG spectrogram reconstruction and oxygen desaturation prediction were removed and replaced with a randomly initialized linear prediction head attached to the RoFormer encoder. Rather than using only the final-layer representation, hidden states from all encoder layers were aggregated through a learnable weighted combination. Specifically, the aggregated representation was computed as 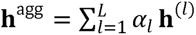, where **h**^(*l*)^ denotes the hidden state from layer *l, L* is the total number of layers, and *α_l_* are learnable aggregation weights initialized such that *α_L_* =1 and *α_1:L-1_* = 0. These weights are optimized during fine-tuning to prioritize task-relevant layers.

During fine-tuning, the model was trained on eight NVIDIA H20 GPUs using Adam with the same batch size and learning rate as used during pretraining. Training was conducted for up to 10 epochs with early stopping based on validation performance.

### Downstream Evaluation Details

#### Baseline methods for comparison

To evaluate the downstream performance of SleepFounder, we established a set of baseline models, including an adapted contrastive pretraining baseline derived from SleepFM**^Error!^ ^Reference^ ^source^ ^not^ ^found.^**(SleepFM-CR), a randomly initialized version of SleepFounder (SleepFounder-Scratch), and conventional task-specific models for sleep staging and OSA detection. For fair comparison, all baselines were adapted to the unified 4 Hz respiratory and heartbeat signals used in this study. Because the original SleepFM framework was designed for multimodal PSG pretraining using cross-modal contrastive learning across channels such as EEG, ECG, electromyography, and respiratory signals, it could not be directly applied to our cardiorespiratory input setting. We therefore implemented SleepFM-CR following the SleepFM paradigm. SleepFM-CR adopts the same architecture and training framework as SleepFM but is pretrained on the cardiorespiratory signals and pretraining corpus used in this study. The comparison with SleepFM-CR was designed to isolate the contribution of our physiology-inspired pretraining strategy relative to a representative contrastive representation learning approach. In addition, SleepFounder-Scratch was used to quantify the contribution of large-scale pretraining beyond supervised task-specific learning. Detailed descriptions and references for baseline methods are provided in Supplementary Methods, Section 4.

#### Task design and evaluation metrics

We evaluated SleepFounder across six categories of downstream tasks, including conventional sleep analysis (five-class sleep staging and moderate-to-severe OSA detection, defined as AHI ≥ 15), demographic profiling (age prediction and sex classification), multi-organ disease detection, and future disease prediction. For most tasks, the model directly generated predictions corresponding to the target outcome. The exception was OSA detection, in which respiratory event probabilities were first estimated at 1-second resolution. A probability threshold optimized on the validation set was then applied to generate binary event labels, and consecutive abnormal segments were merged into apnea or hypopnea events according to duration criteria. The AHI was subsequently computed as the number of detected apnea or hypopnea events per hour of sleep and used to determine OSA status.

For multi-organ disease detection and future disease prediction, outcome labels were derived from International Classification of Diseases (ICD)-coded diagnoses extracted from hospital EHRs and mapped to phecodes for analysis. Participants were included if their EHR records spanned at least one year before and one year after the sleep recording date, ensuring adequate ascertainment around the label-defining window. For disease detection, positive cases were defined as participants with the target diagnosis recorded within ±1 year of the PSG date, whereas negative cases had no record of the target disease throughout their entire EHR history. Participants with diagnoses recorded only outside the ±1-year window were excluded because their disease status at the time of sleep recording could not be determined with sufficient confidence. For future disease prediction, following common practice for new-onset definition, positive cases were defined as participants without prior disease history before PSG (or within 7 days after PSG) who subsequently developed the condition during follow-up, whereas negative cases had no record of the disease at any time. Participants with pre-existing disease were excluded. To reduce trivial prediction driven primarily by demographic characteristics rather than physiological information, sex-specific and pregnancy-related phecodes were manually excluded. Phecodes with a prevalence below 1.5% in the fine-tuning set were also excluded. In total, 1,097 phecodes spanning 16 phecode chapters were evaluated for both disease detection and future disease prediction across all participating centers.

Task-specific optimization objectives were selected according to the prediction target. Sleep staging and sex classification were formulated as classification tasks and optimized using cross-entropy loss, whereas age prediction and OSA estimation were formulated as regression tasks and optimized using mean absolute error (MAE). For multi-organ disease detection, all phecode labels were modeled jointly within a multi-label classification framework and optimized using the mean binary cross-entropy loss across phecodes. Future disease prediction was formulated as a multi-task survival prediction problem, with all phecode-specific survival objectives optimized jointly using the mean Cox proportional hazards loss^Error! Reference source not found.^.

Model performance was evaluated using task-appropriate metrics. For the pretext tasks, performance was assessed using the Pearson correlation coefficient and MAE. Sleep staging performance was evaluated using Cohen’s kappa, which measures agreement between predicted and reference labels while accounting for chance agreement. For OSA detection, performance was quantified using AUROC based on the ranking of predicted AHI values, where an AUROC of 1 indicates perfect discrimination and 0.5 corresponds to random chance. For demographic profiling, age prediction was evaluated using MAE, whereas sex classification was assessed using AUROC. Multi-organ disease detection was evaluated using AUROC. To better characterize performance under class imbalance, we additionally report the area under the Precision-Recall-Gain curve (AUPRG)^39^, a prevalence-adjusted extension of the conventional area under the precision-recall curve. Unlike the conventional area under the precision-recall curve, whose baseline performance equals disease prevalence and therefore varies across phenotypes, AUPRG evaluates performance relative to a no-skill classifier and yields a fixed baseline of 0, enabling more meaningful comparisons across phenotypes with different prevalences. Future disease prediction performance was quantified using the concordance index (C-index).

## Results

### Study Population and Method Overview

SleepFounder was developed using data from 34 large-scale, multi-center, and demographically diverse cohorts in the United States and China. The pretraining and downstream fine-tuning stages included 26 cohorts spanning 60,378 individuals and 668,871 hours of sleep recordings. External evaluation was conducted using five independent PSG cohorts (BIDMC, Stanford, Emory, SSPH, and SRRSH), whereas real-world validation was performed using three multi-center BCG cohorts (n = 2,179) collected using the zero-burden Five Seasons under-pillow BCG monitoring system from IMMHC, SSPH, and WCH. Baseline demographic characteristics of all study populations are summarized in Table 1. Because race classification schemes differed across cohorts, a unified superset of race categories was adopted to preserve source-specific taxonomies while enabling consistent reporting. Summary statistics were calculated at the record level after excluding missing values.

**Table 1.**
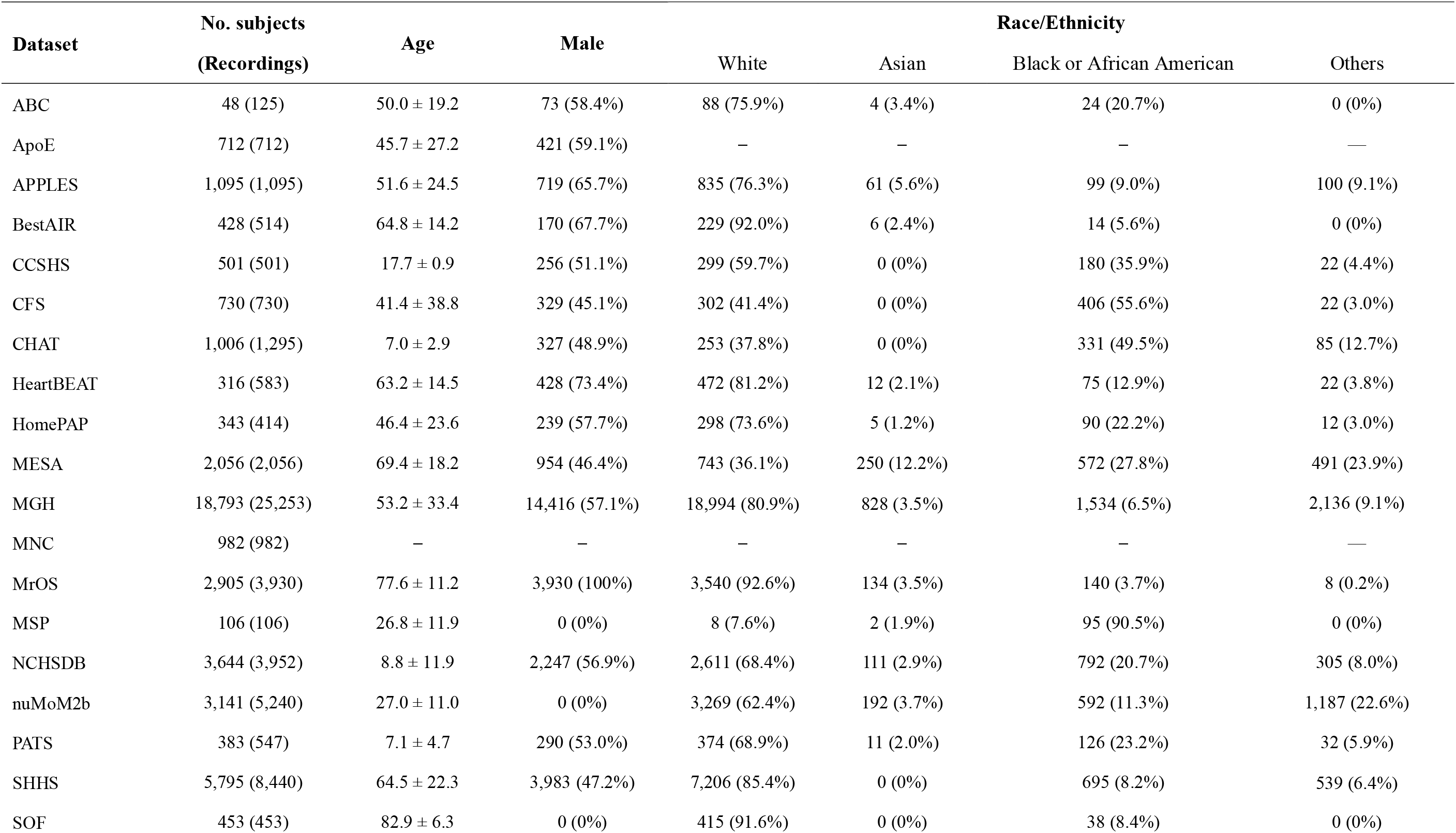
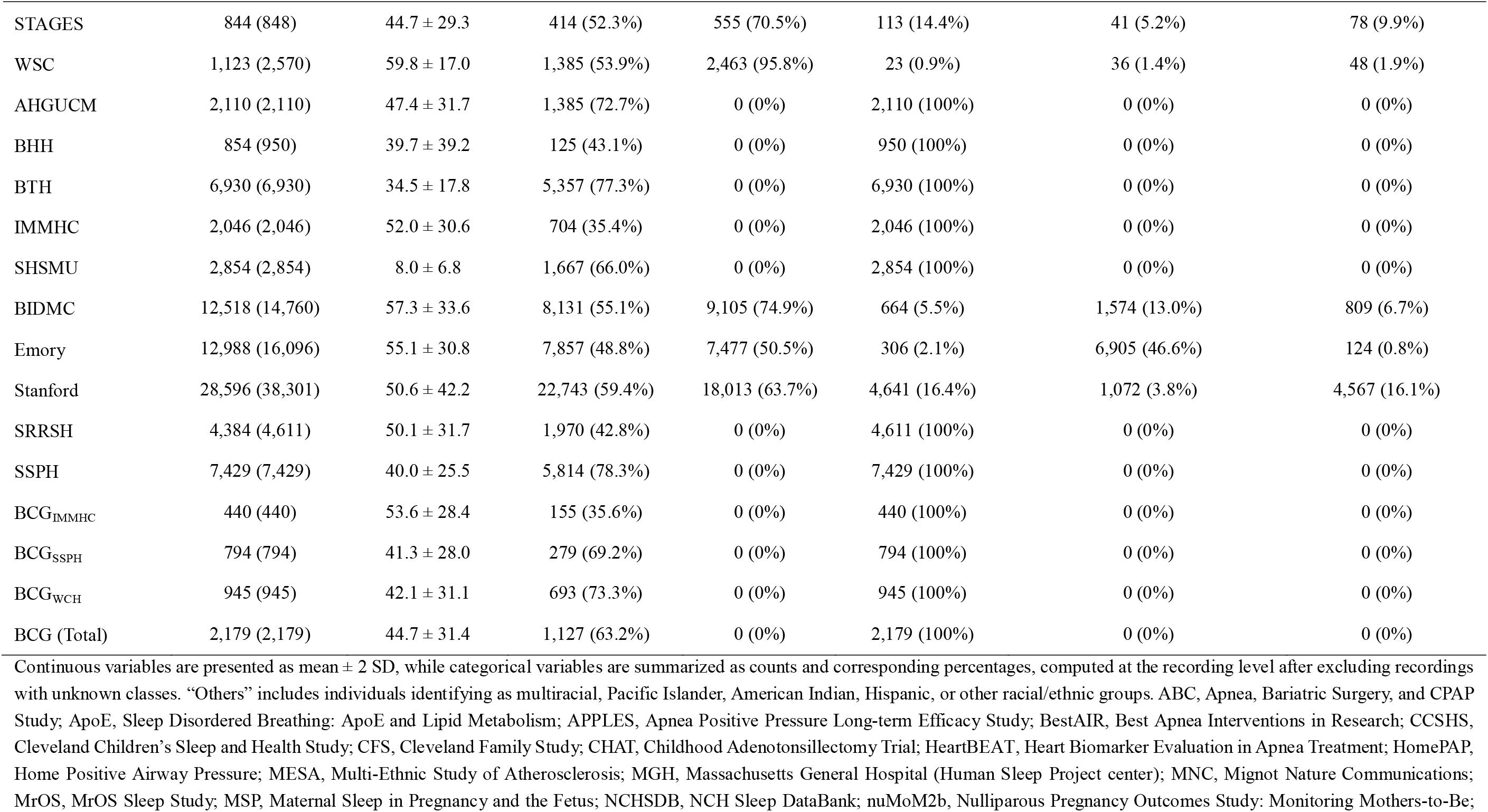
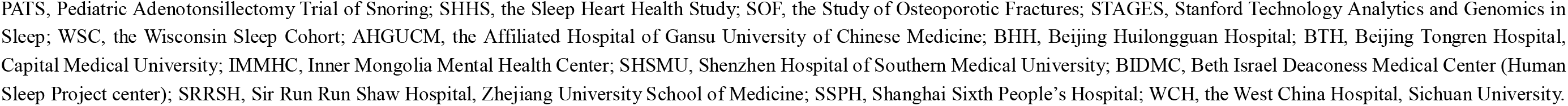
Summary statistics of datasets used for model development.

#### Cardiorespiratory Signals Capture Core Features of Sleep Physiology

SleepFounder was pretrained on physiology-inspired pretext tasks, using cardiorespiratory signals (heartbeat and respiration) to reconstruct EEG spectrograms and predict oxygen desaturation. Qualitatively, the reconstructed outputs closely resembled their target physiological patterns. As shown in representative cases (**Figure 3**a,b), the predicted oxygen desaturation curves accurately tracked the temporal dynamics of measured events across whole nights of sleep, while the reconstructed EEG spectrograms reproduced both the temporal structure and frequency-band composition of the target recordings.

**Figure 3.**
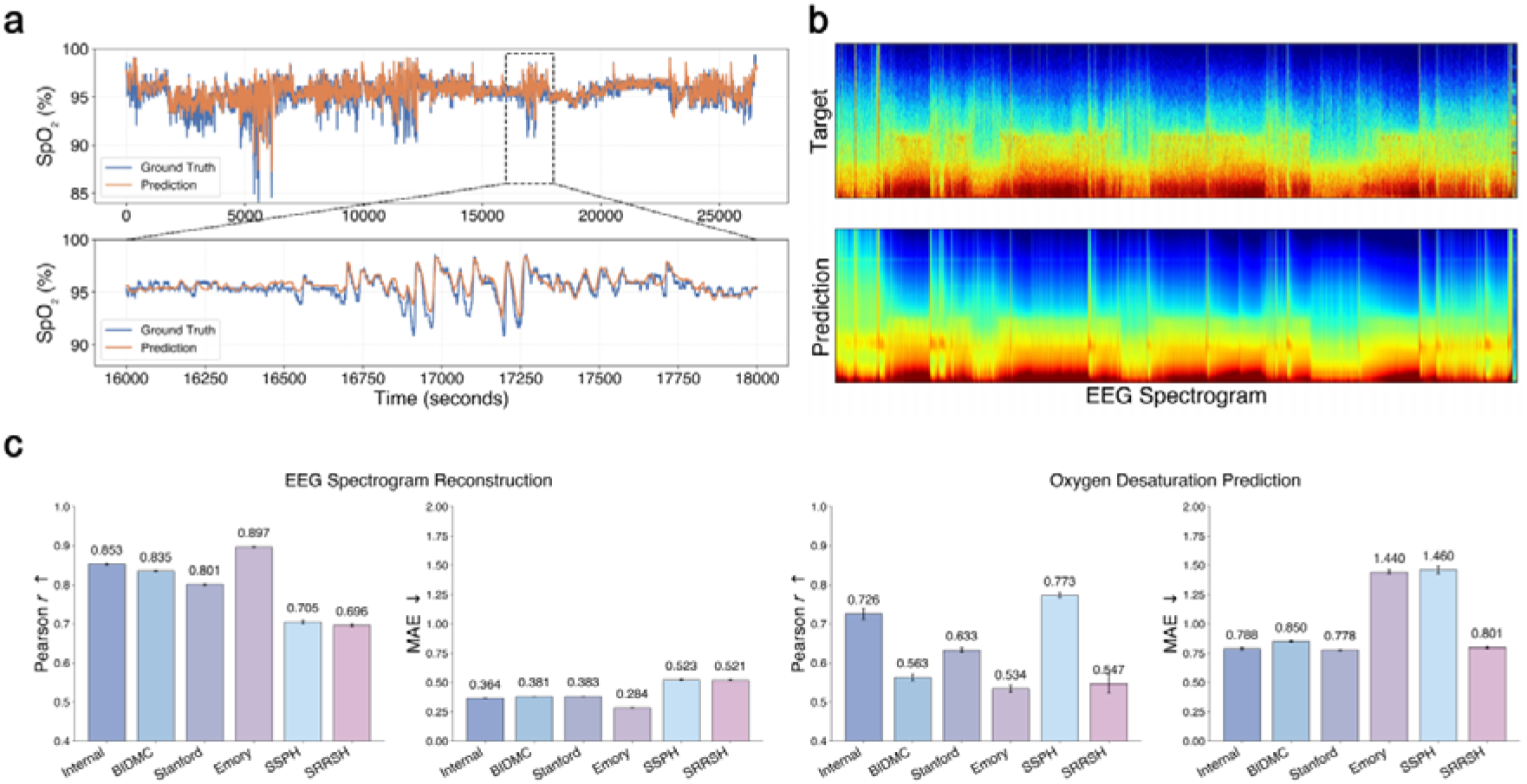
Performance of SleepFounder on physiologically relevant pretext tasks. (a) Example of oxygen desaturation prediction from a single subject’s record, showing close alignment between predicted and target curves. The predicted SpO_2_ curve was obtained by subtracting the predicted desaturation values from the local window maximum baseline derived from the ground-truth SpO_2_. (b) EEG spectrogram reconstruction from the same record, demonstrating that the model can recover sleep-related neural dynamics using only cardiorespiratory signals. (c) Quantitative evaluation of both pretext tasks on the internal dataset and five external cohorts, showing strong and consistent correlations and low mean absolute errors using only cardiorespiratory signals. SpO_2_, blood oxygen saturation; EEG, electroencephalography; Pearson *r*, Pearson correlation coefficient; MAE, mean absolute error; BIDMC, Beth Israel Deaconess Medical Center; SSPH, Shanghai Sixth People’s Hospital; SRRSH, Sir Run Run Shaw Hospital.

Quantitatively, evaluation across the internal dataset and five external cohorts (BIDMC, Stanford, Emory, SSPH, and SRRSH) confirmed that cardiorespiratory signals contain substantial information relevant to neural and oxygenation dynamics during sleep (**Figure 3**c). For EEG spectrogram reconstruction, Pearson correlations reached 0.853 (0.849-0.856) on the internal dataset and remained consistently robust across external cohorts, ranging from 0.696 (0.692-0.700) in SRRSH to 0.897 (0.895-0.898) in Emory. Oxygen desaturation prediction also demonstrated consistent associations, with correlations of 0.726 (0.712-0.739) internally and 0.534-0.773 across external cohorts. These results collectively suggest that overnight cardiorespiratory signals capture key physiological information underlying sleep neurophysiology and oxygenation dynamics.

Beyond pretraining, we benchmarked SleepFounder against task-specific models and input modalities (e.g., full PSG, ECG-only) across downstream tasks (Supplementary Table 8). Despite relying solely on cardiorespiratory signals, SleepFounder achieved performance that was better than, comparable to, or only slightly below that of specialized approaches, demonstrating that fundamental cardiorespiratory activity carries rich physiological information that generalizes across tasks.

#### SleepFounder Demonstrates Superior Performance on Sleep Analysis and Demographic Profiling

SleepFounder was evaluated on two categories of downstream tasks: conventional sleep analysis, including five-class sleep staging and moderate-to-severe OSA detection (hereafter referred to as OSA detection), and demographic profiling, including age prediction and sex classification. Evaluation was conducted on the internal test set and independent external cohorts, including the BIDMC, Stanford, and Emory cohorts in the United States, as well as SSPH and SRRSH in China. Because task labels were not uniformly available across cohorts, OSA detection was additionally evaluated in SSPH, whereas age prediction and sex classification were evaluated in both SSPH and SRRSH.

Across these tasks, SleepFounder achieved strong performance, obtaining the best results in 19 of 21 dataset-task pairs (**Figure 4**a and Supplementary Data 1). In internal validation, SleepFounder achieved a Cohen’s kappa of 0.700 (0.696-0.703) for five-class sleep staging and an AUROC of 0.946 (0.940-0.952) for OSA detection. For demographic profiling, it achieved an MAE of 5.004 (4.896-5.127) years for age prediction and an AUROC of 0.932 (0.925-0.938) for sex classification, with all four internal results representing the best performance among the compared models. In external validation, SleepFounder similarly demonstrated strong generalizability across datasets. It achieved the best results in 15 of 17 external dataset-task pairs and the best averaged performance across all four tasks. Specifically, external averages reached a Cohen’s kappa of 0.651 (0.649-0.652) for sleep staging, an AUROC of 0.912 (0.909-0.915) for OSA detection, an MAE of 7.659 years (7.609-7.708) for age prediction, and an AUROC of 0.889 (0.886-0.891) for sex classification. Compared with a diverse set of baseline models, including conventional task-specific approaches, end-to-end supervised counterparts (SleepFounder-Scratch), and pretrained foundation model (SleepFM-CR), SleepFounder consistently demonstrated superior overall performance, supporting the effectiveness and robustness of its physiology-inspired pretraining strategy.

**Figure 4.**
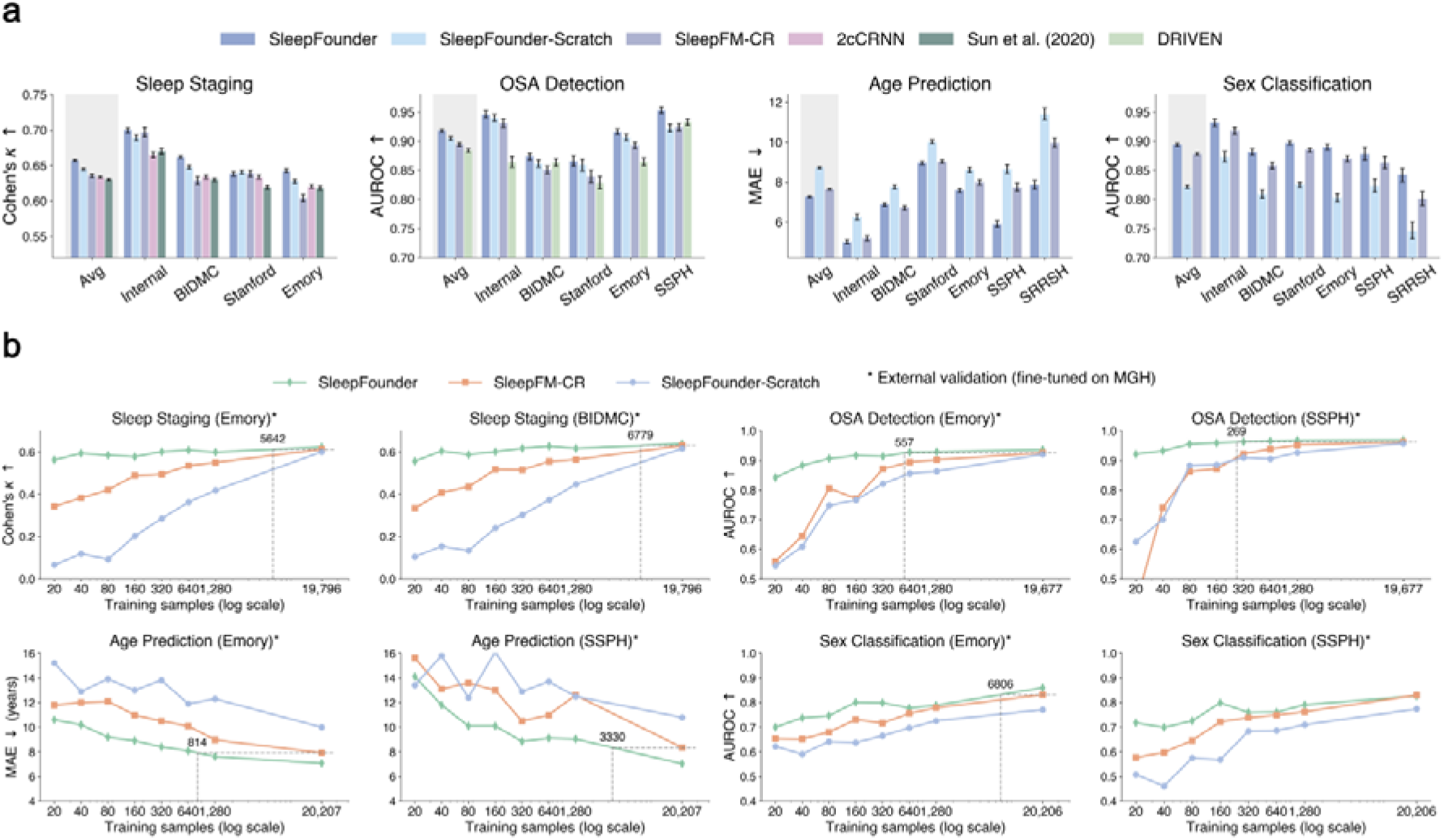
Performance of SleepFounder compared with baseline models across downstream tasks. (a) Internal and external validation results for sleep analysis and demographic profiling tasks, showing that SleepFounder achieved performance comparable to or exceeding that of baseline models in both domains. SleepFM-CR represents a modified version of SleepFM that was adapted to match our input modalities and evaluation protocol, thereby enabling a fair and meaningful comparison with SleepFounder. SleepFounder-Scratch refers to a randomly initialized model with the same architecture as SleepFounder, serving as a control to isolate the contribution of large-scale pretraining. (b) Under limited data conditions, SleepFounder exhibited markedly higher data efficiency, achieving both faster convergence and higher overall performance than baseline models across evaluated tasks. OSA, obstructive sleep apnea; Cohen’s *k*, Cohen’s Kappa; AUROC, area under the receiver operating characteristic curve; MAE, mean absolute error; BIDMC, Beth Israel Deaconess Medical Center; SSPH, Shanghai Sixth People’s Hospital.

We further evaluated the data efficiency of SleepFounder in a cross-cohort generalization setting. Using the MGH data from the fine-tuning set for fine-tuning, we progressively increased the training set size from 20 samples to the full set (approximately 20,000 samples) and assessed performance on downstream tasks across external cohorts: Emory, BIDMC, and SSPH. As shown in **Figure 4**b, SleepFounder exhibited substantially faster convergence than both SleepFM-CR and SleepFounder-Scratch, approaching its full-data performance with considerably fewer training samples. On Emory, for instance, SleepFounder required only 40 training samples (0.2% of the full set) to achieve a Cohen’s kappa of 0.593 on sleep staging, and 80 samples (0.4%) to reach an AUROC of 0.906 on OSA detection, compared with 0.623 and 0.936 under the full training set, respectively.

To further quantify this advantage, we marked the best performance of SleepFM-CR and SleepFounder-Scratch under full training data with horizontal dashed lines in each subplot. As illustrated in **Figure 4**b, the intersection of SleepFounder’s curve with a dashed line corresponds to the estimated number of training samples needed to match that performance level. For sleep staging, SleepFounder required an estimated 5,642 (28.5%) and 6,779 (34.2%) training samples on Emory and BIDMC, respectively, to reach the baseline models’ best performance under full training data. For OSA detection, SleepFounder required an estimated 557 (2.8%) and 269 (1.4%) samples on Emory and SSPH, respectively. Age prediction and sex classification showed similarly efficient data utilization: on Emory, SleepFounder reached the baseline models’ full-data performance with an estimated 814 (4.0%) and 6,806 (33.7%) training samples, respectively. Collectively, these results demonstrate the strong transferability of SleepFounder’s pretrained representations, which enable effective generalization to unseen external populations with limited annotated data.

#### SleepFounder Detects a Broad Spectrum of Diseases from Cardiorespiratory Signals

Sleep is a fundamental physiological state during which multiple organ systems are dynamically regulated and interact, suggesting that overnight cardiorespiratory signals may encode health information extending beyond sleep itself. Motivated by this hypothesis, we evaluated the ability of SleepFounder to detect a broad spectrum of diseases across organ systems through a phenome-wide analysis. Specifically, SleepFounder-derived cardiorespiratory representations were combined with basic demographic features (age and sex), fine-tuned in the MGH cohort across 1,097 ICD-mapped phecodes, and externally evaluated in three independent cohorts (BIDMC, Stanford, and Emory). Performance metrics are reported as average results across the three external cohorts, while full per-cohort and per-phecode results are provided in Supplementary Data 2.

At the phenome-wide level, SleepFounder exhibited broadly distributed disease detection capability (**Figure 5**a). Of the 1,097 phenotypes evaluated, 1,085 (98.9%) achieved an external average AUROC above chance, with an overall median AUROC of 0.652. Detection performance varied systematically across phecode chapters (**Figure 5**b, Supplementary Table 4): the circulatory system showed the highest median AUROC (0.730), consistent with the direct cardiovascular origin of sleep cardiorespiratory signals; endocrine/metabolic, genitourinary and mental disorders formed a second tier, with median AUROCs of 0.687, 0.691 and 0.669, respectively; the lowest medians were observed for sense organs and congenital anomalies (0.617 and 0.616), reflecting their weaker overall coupling to cardiorespiratory physiology. Within this broad distribution, 172 phenotypes (15.7%) achieved an external average AUROC > 0.75, spanning all 16 phecode chapters covered (**Figure 5**a; Supplementary Data 2). The high-performing phenotypes encompassed not only conditions directly related to cardiopulmonary function (Supplementary Data 2), including congestive heart failure (AUROC 0.840, 0.834-0.846), atrial fibrillation (0.835, 0.828-0.842) and respiratory failure (0.822, 0.810-0.833), but also multiple disorders that are not conventionally regarded as sleep-related or cardiorespiratory endpoints, such as dementia (0.850, 0.836-0.862), Parkinson’s disease (0.846, 0.832-0.860), and stage 4 chronic kidney disease (0.829, 0.812-0.846). These results indicate that the health-relevant information carried by sleep cardiorespiratory signals extends substantially beyond sleep-related conditions and conventional cardiopulmonary endpoints. Notably, this broader detection capability was not uniformly observed across all phenotypes. Representative negative-control phenotypes showed lower performance, including conductive hearing loss (0.444) and dermatologic phenotypes such as psoriasis, rash, seborrheic dermatitis, and eczema (AUROC range, 0.543-0.595; Supplementary Data 2). These negative-control results provide additional context for interpreting the specificity of the observed disease associations. **Figure 5**c-d further present subgroup analyses stratified by sex and age. Performance was largely consistent between males and females, with only a modest difference in average AUROC (+0.009 in males). In contrast, disease detection performance was generally higher in younger individuals (<50 years) than in older individuals, with an average AUROC improvement of 0.025.

**Figure 5.**
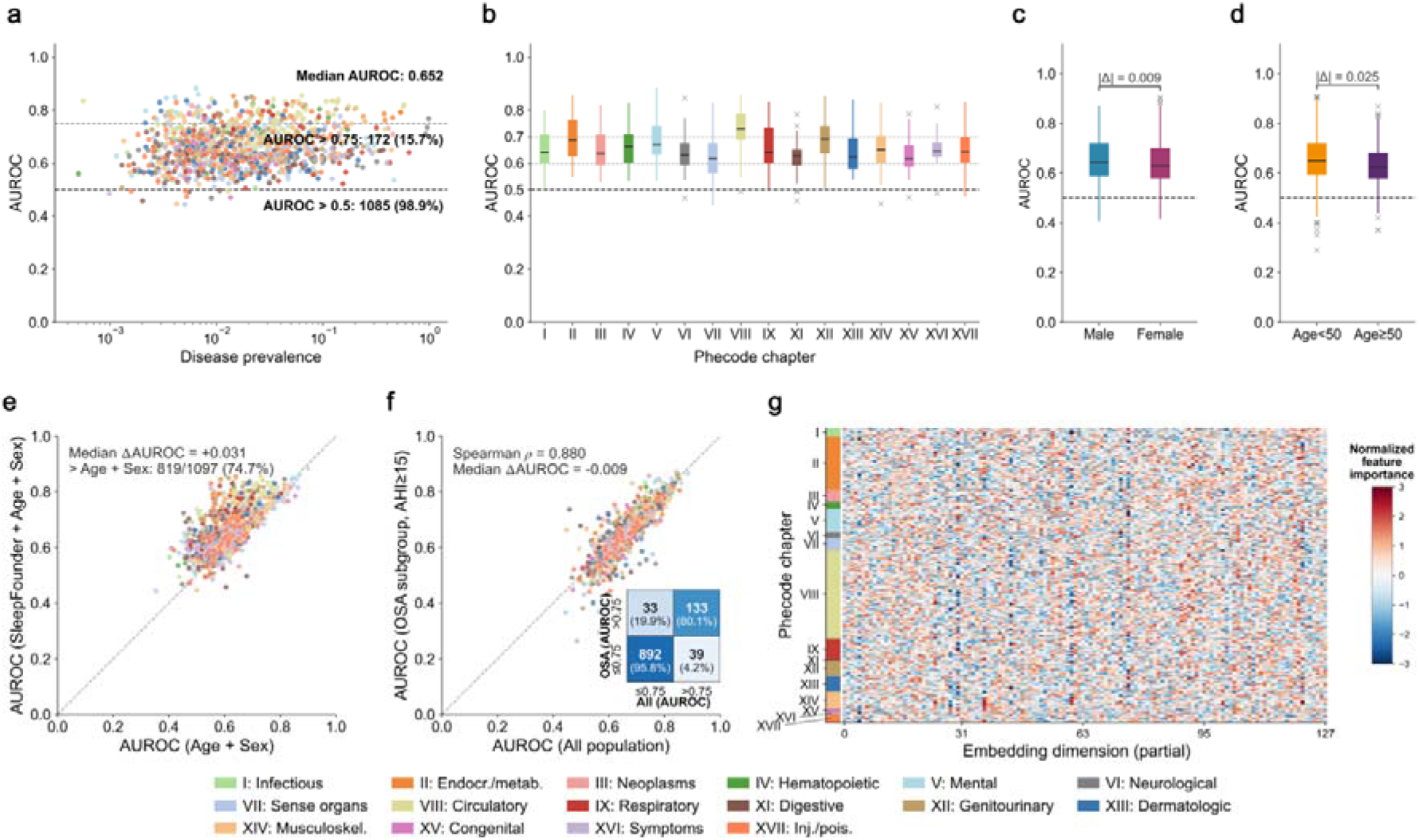
SleepFounder enables broad and phenotype-specific disease detection from sleep cardiorespiratory signals. (a) Phenome-wide scatter plot showing external average AUROC versus disease prevalence for all 1,097 phecodes, with each point colored by phecode chapter. (b) Distribution of external average AUROC stratified by phecode chapter across all 1,097 phecodes. (c) Comparison of external average AUROC distributions between male and female participants, showing broadly consistent disease detection performance across sex subgroups, with males exhibiting an average AUROC 0.009 higher than females. (d) Comparison of external average AUROC distributions between age subgroups (<50 years and ≥50 years), with the younger subgroup exhibiting an average AUROC 0.025 higher than the older subgroup. (e) Comparison of per-phenotype external average AUROC between SleepFounder (cardiorespiratory representations + age + sex) and a demographics-only baseline (age + sex). SleepFounder improved AUROC in 819 of 1,097 phenotypes (74.7%), with a median ΔAUROC of +0.031. (f) Per-phenotype comparison of external average AUROC between the all-population and moderate-to-severe OSA subgroup analyses; the inset confusion matrix summarizes agreement between the two settings based on an AUROC threshold of 0.75. (g) Visualization of the normalized classification-head weight matrix of SleepFounder. Each row corresponds to a disease phenotype and each column corresponds to a feature dimension (first 128 dimensions shown). Color intensity reflects the normalized contribution of each feature dimension to disease prediction, illustrating heterogeneous and phenotype-specific patterns of feature utilization across diseases. AUROC, area under the receiver operating characteristic curve; OSA, obstructive sleep apnea.

To assess whether the broad detection capability of SleepFounder reflects genuine disease-related cardiorespiratory information rather than non-specific shortcuts, we performed three complementary analyses. First, we benchmarked SleepFounder against a matched demographics-only model trained using age and sex under identical fine-tuning and evaluation protocols. Adding SleepFounder representations on top of demographic features improved AUROC in 819 of 1,097 phenotypes (74.7%), with a median ΔAUROC of +0.031 (**Figure 5**e). The improvement was even more pronounced among the high-performing phenotypes, with AUROC improvements, observed in 144 of 172 phenotypes (83.7%) and a median ΔAUROC of +0.078 (Supplementary Figure 8). Second, because OSA itself profoundly alters overnight cardiorespiratory signals and is commin in sleep cohorts, we evaluated SleepFounder in participants with moderate-to-severe OSA. Across the 1,097 phenotypes evaluated in both the all-population and OSA subgroup analyses, disease-level performance remained highly consistent, with a strong correlation between AUROCs obtained in the two settings (Spearman ρ = 0.880, P < 0.001; **Figure 5**f). Finally, we examined whether the broad detection capability reflected a generic comorbidity burden rather than phenotype-specific discriminative patterns. Visualization of the learned disease embedding space for the 172 high-performing phecodes revealed substantial heterogeneity (**Figure 5**g). Consistent with this observation, pairwise cosine similarities among phecode-specific linear classification weight vectors remained uniformly low (median = 0.135; Q1/Q3 = 0.080/0.185), with 95.1% of disease pairs exhibiting |cos| < 0.25. Phenotypes within the same phecode chapter exhibited only slightly higher similarity than the overall similarities across all phenotype pairs (median 0.159 vs. 0.135) (Supplementary Figure 11). Taken together, these analyses suggest that the broad detection capability of SleepFounder cannot be explained solely by demographic risk factors, sleep-disordered breathing, or a non-specific overall disease burden, and instead likely reflects disease-relevant and phenotype-specific information embedded in sleep cardiorespiratory signals.

#### SleepFounder Provides Prognostic Value in Future Disease Prediction

Extending beyond cross-sectional disease detection, we next evaluated whether SleepFounder-derived representations support future disease prediction across multiple organ systems. Prognostic performance was quantified by C-indices averaged across the Stanford, BIDMC, and Emory cohorts, and detailed per-cohort and per-phecode results are provided in Supplementary Data 2.

Consistent with the disease detection setting, prognostic signal remained broadly distributed across phenotypes (**Figure 6**a). Among the 1,097 evaluated phenotypes, 1,081 (98.5%) achieved a C-index above random chance, with an overall median C-index of 0.634; 100 phenotypes (9.1%) further exceeded 0.75. All 16 phecode chapters demonstrated median prognostic performance above the random baseline (**Figure 6**b), with circulatory system phenotypes again showing the strongest performance (median C-index 0.713), followed by genitourinary diseases (0.670), hematopoietic disorders (0.653), endocrine/metabolic diseases (0.652), and mental disorders (0.650), whereas congenital anomalies (0.586) and diseases of the sense organs (0.591) remained at the lower end. **Figure 6**c-d further present subgroup analyses stratified by sex and age. Performance differed only modestly between males and females, with an average C-index difference of 0.014. By comparison, performance was higher among individuals younger than 50 years, with an average C-index 0.025 higher than that observed in older individuals.

**Figure 6.**
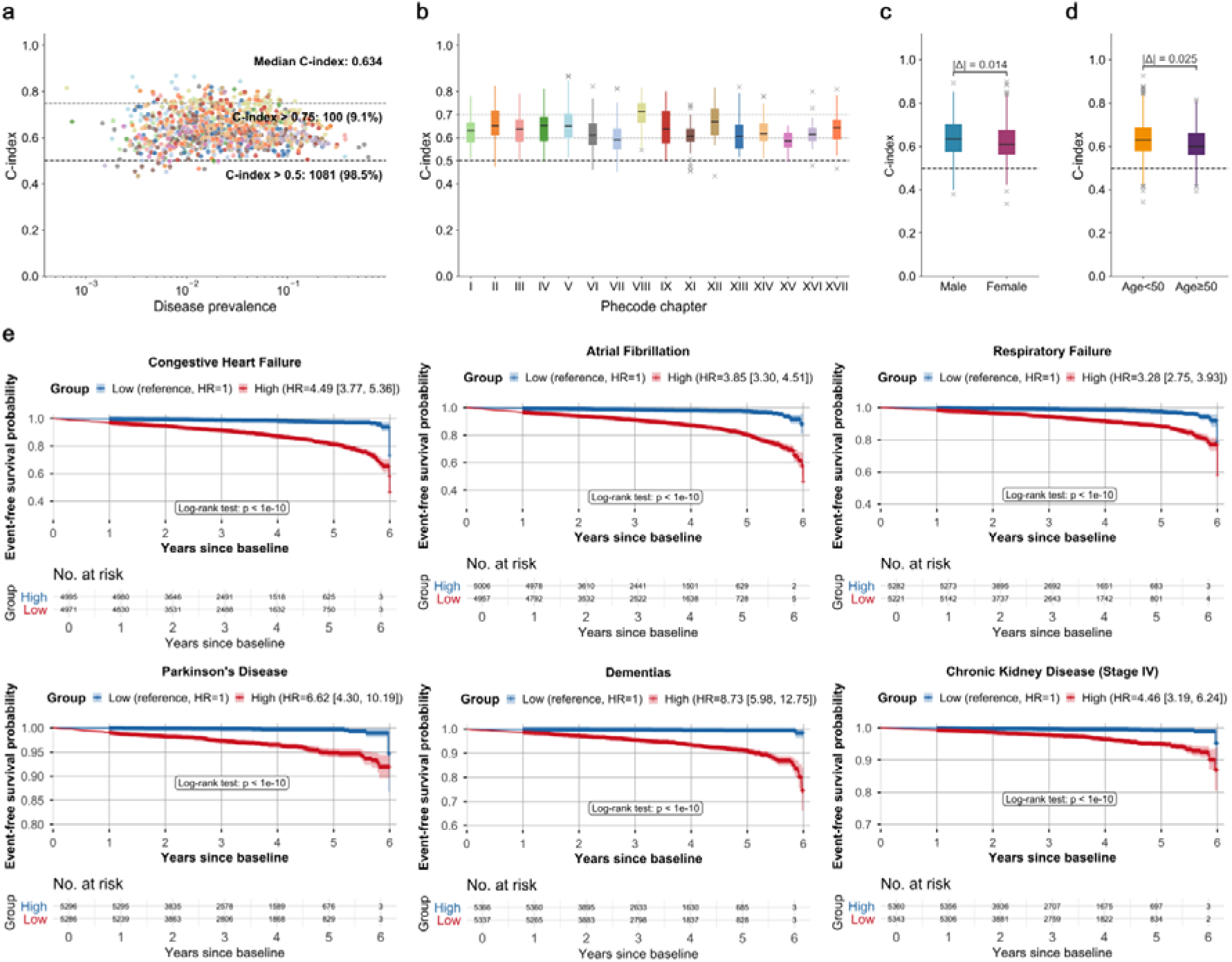
Performance of SleepFounder for future disease prediction. (a) External average C-index for 1,097 incident phecodes plotted against the prevalence of incident occurrences after baseline (log scale). Each point represents one phecode and is colored by phecode chapter. (b) Distribution of external average C-index stratified by phecode chapter across all 1,097 incident phenotypes. (c) Comparison of external average C-index distributions between male and female participants. (d) Comparison of external average C-index distributions between age subgroups (<50 years and _≥_50 years). (e) Kaplan-Meier analyses for representative phenotypes from the external Stanford cohort, including congestive heart failure, atrial fibrillation, respiratory failure, Parkinson’s disease, dementia, and stage 4 chronic kidney disease. Participants were stratified into high-risk and low-risk groups according to the median SleepFounder-predicted risk score (top 50% versus bottom 50% of the population). C-index, concordance index; HR, hazard ratio.

**Figure 6**e presents Kaplan-Meier analyses of several representative phenotypes in the Stanford cohort (see Supplementary Figures 12-13 for the BIDMC and Emory cohorts), including congestive heart failure, atrial fibrillation, respiratory failure, Parkinson’s disease, dementia, and stage 4 chronic kidney disease. Participants were stratified into high-risk and low-risk groups according to the median SleepFounder-predicted risk score (top 50% versus bottom 50% of the population). Across all illustrated phenotypes, the predicted risk scores yielded clear separation of the event-free survival curves during follow-up, with significant differences between the two groups (all log-rank P < 1e-10). The high-risk group consistently exhibited elevated event risk, with hazard ratios ranging from 3.28 for respiratory failure to 8.73 for dementia.

#### SleepFounder Achieves Robust Performance on Real-world BCG Study

SleepFounder was pretrained on cardiorespiratory signals extracted from PSG recordings. To evaluate whether the model itself can be directly applied beyond clinical PSG settings to commonly used home-based devices that capture cardiorespiratory activity, we conducted a real-world study using multi-center BCG signals collected with our previously developed under-pillow BCG monitoring system as a representative example.

For cross-modal agreement assessment, we analyzed recordings where PSG and BCG were acquired simultaneously. As illustrated in a **Figure 7**a-b, BCG-derived respiratory and heartbeat signals showed strong concordance with their PSG counterparts, with respiratory rate achieving near-perfect alignment (Pearson correlation coefficient = 0.94, MAE = 0.29 breaths/min) and heart rate also demonstrating high consistency (Pearson correlation coefficient = 0.91, MAE = 1.84 beats/min). Distribution analyses further confirmed robust cross-modal correlations (**Figure 7**c). These results indicate that the cardiorespiratory signals used to train SleepFounder are reliably captured by BCG devices, thereby supporting the direct applicability of the model in home-based monitoring.

**Figure 7.**
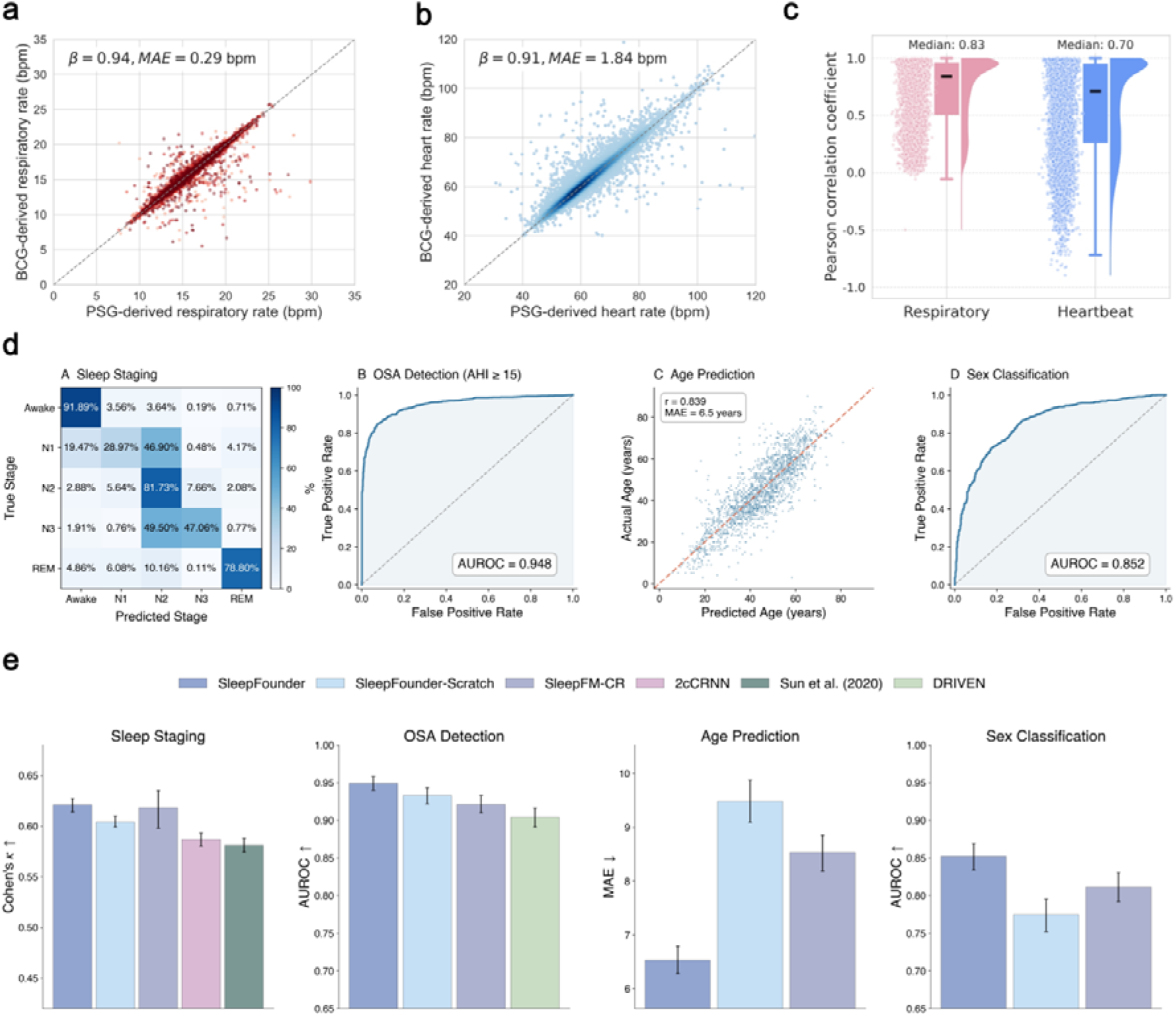
Real-world external validation of SleepFounder on multi-center BCG data. (a-b) Density scatter plots comparing respiratory and heart rates estimated from PSG and BCG signals. (c) Raincloud plots showing the distribution of Pearson correlation coefficients across PSG-BCG pairs for respiratory and heartbeat signals. (d) Performance of SleepFounder across downstream tasks. For sleep staging, a row-normalized confusion matrix illustrates the distribution of predicted sleep stages. The performance on OSA detection and sex classification is summarized using receiver operating characteristic curves, which demonstrate the model’s discriminative performance. For age prediction, a scatter plot presents the relationship between predicted and actual ages, along with the Pearson correlation coefficient (r). (e) Performance of SleepFounder versus baseline models across downstream tasks in the external, multi-center BCG dataset. SleepFM-CR represents a modified version of SleepFM that was adapted to match our input modalities and evaluation protocol, thereby enabling a fair and meaningful comparison with SleepFounder. SleepFounder-Scratch denotes a randomly initialized model with the same architecture as SleepFounder, serving as a control to assess the contribution of large-scale pretraining. PSG, Polysomnography; BCG, ballistocardiography; bpm, breaths/beats per minute; β, Pearson correlation coefficient; MAE, mean absolute error; OSA, obstructive sleep apnea; AHI, apnea-hypopnea index; AUROC, area under the receiver operating characteristic curve; Cohen’s *k*, Cohen’s Kappa.

To further evaluate the real-world applicability of SleepFounder, we performed external validation using BCG datasets collected from three independent clinical centers in China, all of which were excluded from both pretraining and fine-tuning. Remarkably, without any additional adaptation, SleepFounder (fine-tuned in PSG data as in PSG-based downstream evaluation) demonstrated strong and consistent performance across four downstream tasks: a Cohen’s kappa of 0.621 (0.614-0.627) for five-class sleep staging, an AUROC of 0.949 (0.940-0.958) for OSA detection, an MAE of 6.533 years (6.278-6.792) for age prediction, and an AUROC of 0.852 (0.834-0.869) for sex classification (**Figure 7**d). Moreover, when compared with multiple baseline models, SleepFounder consistently matched or outperformed competing approaches across all tasks, underscoring its robustness and direct deployability for real-world home-based sleep monitoring (**Figure 7**e).

## Discussion

In this study, we introduce SleepFounder, a foundation model that leverages widely available cardiorespiratory signals, which are shared physiological modalities in both clinical PSG and home-based systems such as BCG, to enable sleep-centered health assessment. Building on prior PSG-based sleep foundation model research such as SleepFM**^Error!^ ^Reference^ ^source^ ^not^ ^found.^**, SleepFounder shifts the focus toward practical and easily obtainable input signals, thereby offering greater potential for real-world deployment in home-based settings.

Overall, SleepFounder consistently achieved strong performance compared with both self-supervised**^Error!^ ^Reference^ ^source^ ^not^ ^found.^** and task-specific^16,40,41^ baseline models, often matching or surpassing them while using only a fraction of the labeled training data (e.g., ∼2% for OSA detection and ∼30% for sleep staging), highlighting its strong data efficiency (**Figure 4**b). Its performance also compares favorably with previously reported task-specific approaches (Supplementary Table 8). For sleep staging, a prior cardiorespiratory-based study^42^ reported external four-class Cohen’s kappa values of 0.578 (0.576-0.581) on SHHS and 0.643 (0.641-0.645) on MESA, whereas SleepFounder achieved an average Cohen’s kappa of 0.651 [0.649-0.652] across three external cohorts. For age prediction, SleepFounder achieved an MAE of 5.050 (4.770-5.320) years on the SHHS internal test set, compared with previously reported MAEs of 8.2 and 9.4 years for an EEG-based model^43^ on SHHS1 and SHHS2, respectively, and 10.4 and 8.1 years for ECG- and respiration-based models^44^. Although differences in datasets, task definitions, and evaluation protocols preclude direct comparisons, these literature results nevertheless provide a useful point of reference and support the view that overnight cardiorespiratory dynamics contain substantial information for sleep assessment and health profiling.

Given the strong links between sleep physiology and multi-organ health, we evaluated SleepFounder across 1,097 ICD-derived disease phenotypes spanning 16 phecode chapters. SleepFounder demonstrated broadly distributed disease detection capability, with 1,085 of 1,097 phenotypes (98.9%) achieving an external average AUROC above chance and 172 phenotypes exceeding an AUROC of 0.75 across all 16 phecode chapters (Supplementary Data 2). High-performing phenotypes included not only cardiovascular conditions but also disorders such as dementia (0.850, 0.836-0.862), Parkinson’s disease (0.846, 0.832-0.860), and stage 4 chronic kidney disease (0.829, 0.812-0.846), suggesting that the information encoded in overnight cardiorespiratory signals extends well beyond conventional sleep-related endpoints. Notably, performance exhibited a biologically interpretable gradient across disease categories (Supplementary Table 4). Circulatory diseases showed the highest median AUROC (0.730), consistent with the fact that the signals used by SleepFounder directly reflect cardiovascular and respiratory physiology and that cardiovascular disorders substantially alter autonomic regulation, heart rhythm dynamics, and cardiopulmonary coupling during sleep. Genitourinary (median AUROC 0.691) and endocrine/metabolic (median AUROC 0.687) phenotypes also demonstrated relatively strong performance, which is physiologically plausible given the established influences of renal, urologic, and metabolic disorders on fluid balance, blood pressure regulation, autonomic tone, ventilatory control, obesity-related respiratory loading, and sleep-disordered breathing^45,46,47^, all of which can influence overnight respiratory dynamics and heart rate variability. In contrast, phecode chapters with less direct coupling to overnight cardiorespiratory physiology, such as congenital anomalies and diseases of the sense organs, exhibited comparatively lower performance (median AUROCs of 0.616 and 0.617, respectively). Importantly, the observed phenome-wide detection performance was not readily explained by demographic risk factors, sleep-disordered breathing, or a non-specific disease burden. Disease detection performance remained largely preserved in subgroup and baseline analyses (**Figure 5**e-f), while visualization of the normalized classification-head weights (**Figure 5**g) and their pairwise similarities (Supplementary Figure 11) indicated that different disease phenotypes relied on distinct representation patterns rather than a single shared discriminative direction. Nevertheless, it is possible that, for some disease categories, such as musculoskeletal disorders and injuries, model performance partly reflects non-specific downstream effects related to discomfort or functional impairment, rather than disease-specific physiological alterations alone. Collectively, these findings suggest that sleep cardiorespiratory signals contain rich disease-relevant information spanning multiple organ systems and support the use of foundation models as a data-driven framework for systematically characterizing associations between overnight physiology and human health.

In addition to disease detection, SleepFounder demonstrated strong performance for future disease prediction. Among the 1,097 incident phecodes evaluated across three independent external cohorts, 1,081 (98.5%) achieved a C-index above chance and 100 phecodes exceeded a C-index of 0.75. Representative Kaplan-Meier analyses (**Figure 6**e) further demonstrated clear risk stratification across diverse disease categories, with hazard ratios ranging from 3.28 to 8.73 between the upper and lower halves of the SleepFounder-predicted risk distribution. Notably, these prognostic signals were derived from a single night of passively acquired cardiorespiratory recordings. For context, the SCORE2 clinical risk prediction algorithm^48^ achieved a C-index of 0.739 in its derivation cohort, whereas the Framingham Risk Score (Wilson) reported a median C-index of 0.71 for cardiovascular disease prediction in a meta-analysis^49^. In comparison, SleepFounder achieved a median C-index of 0.713 for circulatory-system phenotypes averaged across three independent external cohorts (Supplementary Table 5), suggesting the potential of overnight cardiorespiratory physiology to serve as a scalable, low-burden complement to conventional risk assessment approaches that rely on clinical, laboratory, and demographic variables.

Beyond these findings, external validation using multi-center BCG data collected from three independent institutions in China demonstrated the real-world applicability of SleepFounder for home-based sleep monitoring (**Figure 7**). Across downstream tasks, performance on BCG recordings remained comparable to that observed in PSG-based external validation, despite the model being developed exclusively using PSG-derived cardiorespiratory signals. For example, SleepFounder achieved a Cohen’s kappa of 0.621 [0.614-0.627] for five-class sleep staging and an AUROC of 0.949 [0.940-0.958] for OSA detection on external BCG cohorts, compared with corresponding PSG-based external validation performance of 0.651 [0.649-0.652] and 0.912 [0.909-0.915], respectively. These findings complement recent work such as BCGNet^36^, which demonstrated accurate sleep staging and AHI estimation on the same under-pillow BCG hardware platform using device-specific supervised training. Rather than focusing on device-specific optimization, SleepFounder evaluates whether representations learned from PSG-derived cardiorespiratory signals can generalize beyond conventional PSG acquisition to real-world contactless monitoring settings, with BCG serving as a representative validation platform. These findings suggest that the representations learned by SleepFounder capture physiological information that is largely preserved across sensing modalities rather than being specific to a particular acquisition device. More broadly, the successful transfer from PSG to contactless BCG supports the potential of cardiorespiratory foundation models for scalable, low-cost, and contact-free sleep and health monitoring in real-world settings.

Despite these promising results, several limitations warrant consideration. First, although SleepFounder underwent extensive validation across multiple large-scale PSG cohorts from both the United States and China, as well as real-world BCG recordings collected from three independent clinical centers, pretraining relied exclusively on PSG data, and the real-world BCG validation was conducted using a single under-pillow monitoring system. While these experiments provide encouraging evidence of cross-cohort and cross-modality generalizability, further evaluation across broader populations, more diverse real-world environments, independent contactless sensing platforms, and other wearable monitoring devices will be important to more comprehensively establish the model’s real-world applicability and device generalizability. Second, comparisons with existing foundation models relied on adapted implementations (e.g., SleepFM-CR), as no published foundation model is currently designed for cardiorespiratory-only inputs. Although every effort was made to ensure fair comparison, adapted implementations may not fully reproduce the performance of the original models in their intended settings. Third, although multi-organ disease detection and future disease prediction were evaluated across multiple independent HSP centers, these analyses were conducted exclusively in U.S.-based cohorts and have not yet been validated in more diverse international populations or real-world contactless monitoring settings. In addition, while SleepFounder demonstrated strong transferability to contactless BCG recordings for conventional downstream tasks, disease detection and prediction have not yet been evaluated on contactless devices due to the current lack of corresponding disease outcome labels. Future studies involving broader populations, additional healthcare systems, and real-world cohorts with linked clinical outcomes will be important to further assess the generalizability and clinical utility of SleepFounder.

In conclusion, SleepFounder demonstrates that cardiorespiratory-based foundation models can enable accurate, scalable, and zero-burden sleep monitoring. Built upon large-scale, multi-ethnic PSG pretraining and validated across both clinical PSG and BCG datasets, the model achieved robust and generalizable performance across a spectrum of sleep and multi-organ health tasks. By integrating accessible cardiorespiratory signals with physiology-informed pretraining, SleepFounder establishes a scalable framework for continuous, real-world, and individualized AI-driven sleep and health assessment. These findings extend foundation-model applications beyond traditional PSG and introduce a new paradigm for precision preventive medicine, in which passive physiological monitoring enables early risk detection, targeted intervention, and proactive health management in out-of-clinic settings.

## Supporting information

Supplementary meterial

## Data Availability

The Human Sleep Project (HSP) and the National Sleep Research Resource (NSRR) datasets analyzed in this study are publicly available. The datasets collected in China are not publicly available due to privacy and regulatory restrictions but may be made available from the corresponding author upon reasonable request.

https://bdsp.io/content/hsp/2.0/

https://sleepdata.org/

## Acknowledgments

This study was conducted as part of the AISleep 365 Consortium, a multi-center harmonization initiative uniting hospital-based PSG data across China to enable AI-driven and home-based sleep health monitoring. We thank the participating hospitals and investigators of the AISleep 365 Consortium for data sharing and collaboration. This work is supported by the Ministry of Science and Technology of China STI2030-Major Projects (No.2021ZD0201900, 2021ZD0201902), and the National Natural Science Foundation of China (62102008).

The authors gratefully acknowledge the National Sleep Research Resource (NSRR; sleepdata.org), funded by the National Heart, Lung, and Blood Institute (NHLBI), for providing access to de-identified clinical data that supported the development and evaluation of our work. We also thank the Brain Data Science Platform (BDSP) for access to the HSP dataset, a growing collection of clinical polysomnography recordings from Massachusetts General Hospital.

