## Supplementary meterial for "A Zero-Burden Sleep Foundation Model Built on Cardiorespiratory Signals from 1,400,000+ Hours of Multi-Ethnic Sleep Recordings"

**Supplementary Methods**

**Section 1. Dataset descriptions**

1.1 **Datasets for model pretraining and downstream fine-tuning**

The datasets used for model pretraining and downstream fine-tuning comprised 60,378 participants from 26 cohorts in the United States and China. The U.S. component comprised 45,584 participants from 21 datasets, including the Massachusetts General Hospital (MGH) cohort of the Human Sleep Project (HSP) and 20 datasets from the National Sleep Research Resource (NSRR). The Chinese component comprised 14,794 participants from five private clinical cohorts: the Affiliated Hospital of Gansu University of Chinese Medicine (AHGUCM), Beijing Huilongguan Hospital (BHH), Inner Mongolia Mental Health Center (IMMHC), Shenzhen Hospital of Southern Medical University (SHSMU), and Beijing Tongren Hospital, Capital Medical University (BTH).

**MGH**: MGH is one of the participating centers in the HSP dataset ([https://bdsp.io/content/hsp/2.0/](https://bdsp.io/content/hsp/2.0/" \t "/Users/nieguangkun/Documents\\x/_new)), a large-scale sleep research resource comprising overnight polysomnography (PSG) recordings acquired according to American Academy of Sleep Medicine standards. Available signals include electroencephalography (EEG), electrooculography (EOG), chin electromyography (EMG), respiratory effort, airflow, oxygen saturation, and electrocardiography (ECG). In this study, 18,793 individuals with 25,253 PSG recordings were included, with linked electronic health records (EHRs) enabling access to corresponding International Classification of Diseases (ICD)-coded diagnoses.

**NSRR**: The NSRR (<https://sleepdata.org/>) hosts polysomnography (PSG) datasets together with demographic and clinical information from multiple cohorts. For model pretraining, we utilized 20 NSRR datasets, including the Apnea, Bariatric Surgery, and CPAP Study (ABC); Sleep Disordered Breathing: ApoE and Lipid Metabolism (ApoE); Apnea Positive Pressure Long-term Efficacy Study (APPLES); Best Apnea Interventions in Research (BestAIR); Cleveland Children’s Sleep and Health Study (CCSHS); Cleveland Family Study (CFS); Childhood Adenotonsillectomy Trial (CHAT); Heart Biomarker Evaluation in Apnea Treatment (HeartBEAT); Home Positive Airway Pressure (HomePAP); Multi-Ethnic Study of Atherosclerosis (MESA); Mignot Nature Communications (MNC); MrOS Sleep Study (MrOS); Maternal Sleep in Pregnancy and the Fetus (MSP); NCH Sleep DataBank (NCHSDB); Nulliparous Pregnancy Outcomes Study: Monitoring Mothers-to-Be (nuMoM2b); Pediatric Adenotonsillectomy Trial of Snoring (PATS); Sleep Heart Health Study (SHHS); Study of Osteoporotic Fractures (SOF); Stanford Technology Analytics and Genomics in Sleep (STAGES); and Wisconsin Sleep Cohort (WSC). Collectively, these datasets comprised 26,791 individuals and a total of 35,125 PSG recordings.

**AHGUCM**: The AHGUCM dataset was collected at the Sleep Disorders Department of the Affiliated Hospital of Gansu University of Chinese Medicine between 2017 and 2025, comprising overnight PSG recordings from 2,110 patients. Most patients in this cohort were diagnosed with obstructive sleep apnea (OSA) and insomnia.

**BHH**: The BHH dataset was acquired at the Sleep Disorders Center of Beijing Huilongguan Hospital between 2022 and 2025. It consists of overnight PSG recordings from 854 patients with psychiatric disorders, including insomnia, depression, and anxiety.

**IMMHC**: The IMMHC dataset comprises overnight PSG recordings from 2,046 participants collected between 2019 and 2024 at the Sleep Medicine Center of Inner Mongolia Mental Health Center. Participants were patients with psychiatric disorders and comorbid OSA.

**SHSMU**: The SHSMU dataset comprises 2,854 overnight PSG recordings collected from 2017 to 2024 at the Department of Pediatric Otorhinolaryngology, Shenzhen Hospital of Southern Medical University. This cohort specifically consists of children, with a mean age of 7.97 years.

**BTH**: The BTH dataset includes overnight PSG recordings from 6,930 participants collected between January 2017 and December 2024 at the Department of Otorhinolaryngology, Beijing Tongren Hospital. Most participants were patients referred for evaluation of suspected OSA.

1.2 **Datasets for model evaluation**

To evaluate SleepFounder, we utilized both PSG- and ballistocardiography (BCG)-based datasets. PSG evaluation included three HSP cohorts (Stanford, Emory, and Beth Israel Deaconess Medical Center (BIDMC)) as well as two independent clinical cohorts from China, Sir Run Run Shaw Hospital, Zhejiang University School of Medicine (SRRSH) and Shanghai Sixth People’s Hospital (SSPH). In addition, real-world validation was conducted using multi-center BCG datasets collected in parallel with clinical PSG recordings at SSPH, West China Hospital, and IMMHC.

**Stanford**: Stanford is a participating center in the HSP dataset. In this study, 28,596 individuals with 38,301 PSG recordings were included, with linked EHRs providing corresponding ICD-coded diagnoses.

**Emory**: Emory is a participating center in the HSP dataset. In this study, 12,988 individuals with 16,096 PSG recordings were included, with linked EHRs providing corresponding ICD-coded diagnoses.

**BIDMC**: BIDMC is a participating center in the HSP dataset. In this study, 12,518 individuals with 14,760 PSG recordings were included, with linked EHRs providing corresponding ICD-coded diagnoses.

**SRRSH**: The SRRSH dataset refers to data sourced from the Department of Psychiatry at Zhejiang University School of Medicine’s Sir Run Run Shaw Hospital, collected between 2018 and 2023, and includes 4,611 usable recordings from 4,384 paticipants. There is no annotation for sleep stages and apnea-hyponea index (AHI) events for this dataset.

**SSPH**: The SSPH dataset represents data collected between October 2010 and July 2023 from the Department of Otorhinolaryngology at Shanghai Sixth People’s Hospital, consisting of 7,429 PSG recordings from 7,429 participants. It focuses on patients with OSA and includes annotations for sleep stages and respiratory events.

**BCG**: The BCG dataset comprises a total of 2,179 PSG recordings from 2,179 participants, including 945 from the Sleep Medicine Center of West China Hospital (collected between 2024 and 2025), 794 from SSPH (collected from 2023 to 2025), and 440 from IMMHC (collected in 2025). For all recordings, PSG and BCG signals were acquired simultaneously, with respiratory events and sleep stages annotated based on the PSG data. Importantly, this dataset does not overlap in patient population with the previously described IMMHC and SSPH datasets.

**Section 2. Data Preprocessing**

**Heartbeat Component** To capture heartbeat dynamics, we adopted inter-beat intervals (IBIs) as part of the model input. This choice allows us to preserve essential temporal information while significantly reducing the input signal’s sampling frequency. For ECG signals, we first denoised the data and detected R peaks using NeuroKit2^1^, from which the RR intervals were extracted as the IBIs. For BCG signals, we firstly detected J peaks^2^, and the resulting JJ intervals were used as the IBIs. Once the raw IBIs were obtained, we applied the *get_nn_intervals* function from HRVAnalysis^3^ to suppress outliers and artifacts, mitigating their impact on the model. Finally, we linearly interpolated the cleaned IBI sequence at a fixed sampling rate of 4 Hz to generate a continuous heartbeat signal.

**Respiratory Component** The respiratory signal utilized in this work was extracted from the abdominal respiratory effort (ABD effort) in PSG recordings and from raw BCG signals. To ensure consistency and quality across modalities, a four-step preprocessing pipeline was employed, consisting of bandpass filtering, temporal resampling, integration (if necessary), and normalization. First, a third-order Bessel bandpass filter^4^ with a passband of $\frac{1}{10}$ Hz to$\frac{1}{3}$ Hz was applied to suppress high-frequency noise and low-frequency drift. Next, all respiratory signals were resampled to 4 Hz to ensure alignment in temporal granularity. Due to the nature of BCG signals capturing derivative-like respiratory motion, a temporal integration step was applied to approximate respiratory volume and reduce phase mismatch relative to the ABD effort signal. This adjustment is not required for ABD-derived signals, which already represent respiratory amplitude. In the final step, all processed respiratory signals were standardized using z-score normalization to address inter-sensor variability and unify amplitude scales across modalities.

**Section 3. Implementation Details of Reconstruction-Based Pretraining Tasks**

**Electroencephalography spectrogram reconstruction**: The selection of EEG channels was prioritized based on their sensitivity to key sleep-related features such as sleep spindles, slow waves, and rapid eye movement activity. Specifically, we selected channels in the following order: C3/C4 > F3/F4 > O1/O2. The EEG signals corresponding to these channels were preprocessed into spectrograms using the Python LSPOpt package (<https://github.com/hbldh/lspopt>), which provides an optimal Wigner spectrum estimate for locally stationary processes (LSPs). To ensure consistent feature scaling, we applied log transformation and normalization. Specifically, the spectrogram was normalized by the standard deviation and mean of the spectrogram values across frequency bands and time segments. Additionally, linear interpolation was applied to adjust the frequency resolution between 0.3 Hz and 35 Hz. The entire preprocessing process is described by:

$$S=\frac{10\log_{10} (\mathrm{LSPOpt}(x)+1e^{-13})-\mu}{\sigma}$$

where $x$ represents the raw EEG signal, $\mathrm{LSPOpt}(x)$ denotes the locally stationary process estimation of $x$ using the Wigner spectrum, $\mu$ is the mean, and $\sigma$ is the standard deviation of the spectrogram values across frequency bands and time segments. The spectrogram values were then linearly interpolated between the frequencies 0.3Hz and 35Hz, and the logarithmic transformation was applied to scale the spectrogram values.

**Peripheral capillary oxygen desaturation prediction**: SpO_2_ represents the percentage of oxygen-saturated hemoglobin in the blood and reflects the efficiency of oxygen transport from the lungs to peripheral tissues, which is essential for maintaining normal physiological function. After resampling the SpO2 signal to a 1 Hz sampling frequency, the validity of each signal point was assessed by comparing it with its neighboring points. Points exhibiting a drop rate greater than 5 or a recovery rate greater than 10 were considered unreliable and replaced with a NaN value. If more than 40% of the data points within a given night were deemed erroneous, the entire SpO2 signal for that night was discarded. Instead of using the raw SpO2 signal, we computed oxygen desaturation as the learning target. Oxygen desaturation refers to the decrease in oxygen saturation relative to the highest local peak, capturing episodes of hypoxemia. This approach mitigates the effects of individual baseline differences in SpO2 levels, offering a more generalized representation of oxygen fluctuations. The local maximum used for this calculation was determined within a window of 100 seconds.

**Section 4. Baseline Methods for Comparison**

**Self-supervised pretrained models**: We included SleepFM^5^ as a representative multimodal, self-supervised baseline (SleepFM-CR). SleepFM employs deep encoders with a contrastive learning objective across multiple physiological modalities. For a fair comparison with SleepFounder, we adapted SleepFM by adding a respiration-heartbeat branch consistent with our input setting and architecture. During fine-tuning, only this branch was used to align with SleepFounder’s training protocol. Unlike the original SleepFM, which was pretrained primarily on one large-scale PSG dataset, our adaptation relied on a more heterogeneous collection of datasets. While the overall scale was comparable, the greater heterogeneity and stricter modality requirements of SleepFM limited the inclusion of several portable-device PSG datasets, potentially introducing noise and reducing usable data during pretraining.

**Baseline models for sleep staging**: We reproduced two representative convolutional-recurrent architectures. The first was the two-channel network of Goldammer et al.^6^, which processes 4-Hz interval-derived streams with stacked 1D convolutions followed by a bidirectional LSTM for epoch-wise staging. Since the original inputs were also sampled at 4 Hz, they were directly replaced by the respiration and heartbeat signals used in this work; the only modification was extending the LSTM’s temporal context to full-night sequences (up to 1,535 × 30-s epochs), which improved performance relative to short-window settings. The second baseline was the CNN-LSTM model of Sun et al.^7^, originally designed for high-rate ECG (200 Hz) and abdominal/thoracic respiratory effort (10 Hz). In our adaptation, the abdominal CNN encoder was duplicated to process both the preprocessed respiratory signal and the JJ intervals, with signals upsampled to 10 Hz to match the expected input. The original three-stage training procedure was retained, with the only architectural change being an extension of the LSTM temporal window from 28 × 30-s epochs (14 min) to full-night sequences, consistent with SleepFounder.

**Baseline models for AHI estimation**: We reproduced DRIVEN^8^, a representative deep-learning baseline for AHI estimation that combines 2D convolutional neural networks with LightGBM. In the original implementation, DRIVEN jointly performed respiratory-event detection and sleep/wake classification to derive total sleep time, but for a fair comparison with SleepFounder, ground-truth sleep labels were used instead. We note that the omission of oximetry, which is included in the original model, may modestly reduce performance relative to the reported results.

**Section 5. Model Architecture of SleepFounder**

SleepFounder adopts a hybrid architecture (~26.6M parameters) consisting of dual ResNet encoders for modality-specific feature extraction and a RoFormer backbone for long-range temporal modeling (Supplementary Figure 1). The dual ResNet encoders contain approximately 7.7M parameters (3.84M parameters per branch), while the transformer backbone contains approximately 18.9M parameters. Detailed architectural specifications are provided in Supplementary Tables 1-2.

Respiratory and heartbeat signals are processed independently through two structurally identical but separately parameterized ResNet branches, each receiving the full-night continuous physiological waveform as input. Each branch begins with a convolutional stem consisting of a one-dimensional convolutional layer, batch normalization, ReLU activation, and max pooling, resulting in an initial four-fold reduction in temporal resolution. The extracted features are subsequently propagated through four residual stages, each comprising two residual blocks with convolutional layers, batch normalization, ReLU activation, and residual shortcut connections. Feature dimensionality progressively increases across stages (64 → 64 → 128 → 256 → 512), while stage-wise strides of 1, 2, 3, and 5 progressively downsample the temporal axis. The modality-specific latent representations are subsequently fused through element-wise summation to form a unified cardiopulmonary representation. This parameter-free fusion strategy preserves temporal correspondence between modalities while enabling each branch to specialize in modality-specific physiological dynamics.

To capture long-range temporal dependencies across overnight recordings, the fused token sequence is processed using a RoFormer encoder. A learnable [CLS] token is prepended to aggregate global contextual information across the sequence. The RoFormer backbone comprises six transformer layers, each configured with eight self-attention heads (head dimension = 64), a hidden dimension of 512, and a feedforward dimension of 2,048. Rotary positional embeddings encode relative positional information through rotational transformations applied to query and key vectors, enabling efficient modeling of variable-length recordings without fixed positional constraints.

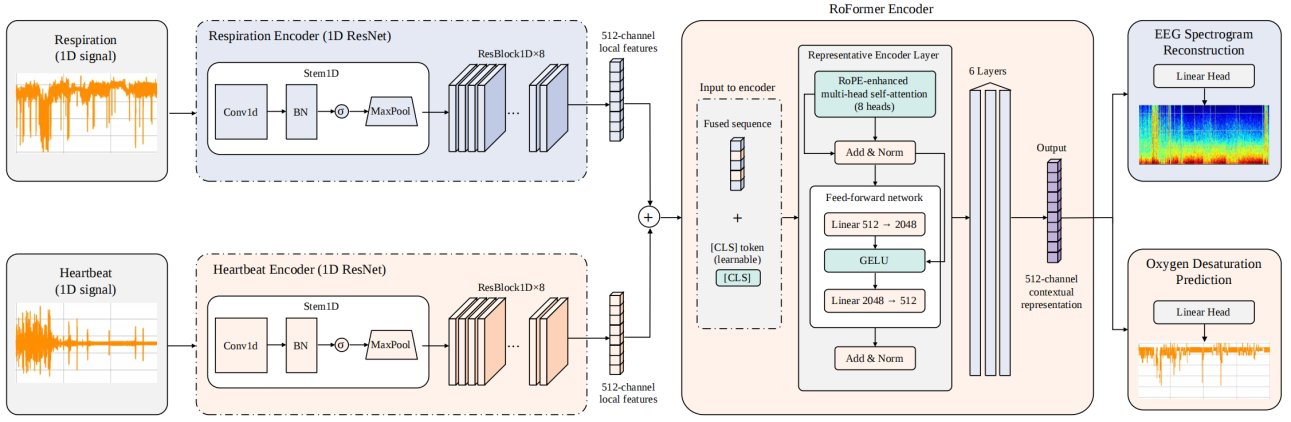

**Supplementary** **Figure 1. Schematic overview of the SleepFounder architecture.** SleepFounder consists of dual ResNet encoders for modality-specific cardiorespiratory feature extraction, followed by a RoFormer encoder for global temporal modeling of overnight cardiorespiratory signals. RoPE, rotary positional embedding.

**Supplementary Table 1: Architecture and parameter summary of SleepFounder.** Output shapes are expressed as [B, C, L] for convolutional layers and [B, L, d] for transformer layers, where B is the batch size, C is the number of channels, L is the temporal length, and d = 512 is the hidden dimension. T = L/120 denotes the number of output tokens (e.g., T = 960 for an 8-hour recording at 4 Hz).

| ***Component*** | ***Input dim*** | ***Output dim*** | ***Output Shape*** | ***#Parameters*** |
| --- | --- | --- | --- | --- |
| **ResNet Branch (×2, independent)** |  |  |  |  |
| Stem | 1 | 64 | [B, 64, L/4] | 576 |
| ResStage (stride=1) | 64 | 64 | [B, 64, L/4] | 49,664 |
| ResStage (stride=2) | 64 | 128 | [B, 128, L/8] | 181,504 |
| ResStage (stride=3) | 128 | 256 | [B, 256, L/24] | 723,456 |
| ResStage (stride=5) | 256 | 512 | [B, 512, L/120] | 2,888,704 |
| Subtotal (per branch) | — | — | [B, 512, T] | 3,843,904 |
| **Modality Fusion (element-wise sum)** | 512 | 512 | [B, 512, T] | 0 |
| **RoFormer Encoder** |  |  |  |  |
| [CLS] token prepend | — | — | [B, T+1, 512] | 512 |
| TransformerLayer × 6 | 512 | 512 | [B, T+1, 512] | 18,914,304 |
| Subtotal (RoFormer) | — | — | [B, T+1, 512] | 18,915,840 |
| **Total** | **—** | **—** | **[B, T+1, 512]** | **26,603,648** |

**Supplementary Table 2. Module definitions of SleepFounder.** Each ResStage consists of two residual blocks. C_in_ and C_out_ denote the input and output channel dimensions, respectively; k denotes kernel size; s denotes stride; and p denotes padding size. d=512 denotes the hidden dimension, and d_k_ denotes the dimension of each attention head. RoPE, rotary positional embedding.

| ***Module*** | ***Definition*** |
| --- | --- |
| **Stem (C_in_, C_out_)** | Conv1d(C_in_, C_out_, k=7, s=2, p=3)  BatchNorm1d(C_out_)  ReLU  MaxPool1d(k=3, s=2, p=1) |
| **ResStage (C_in_, C_out_, s)** | ResBlock(C_in_, C_out_, s=s)  ResBlock(C_out_, C_out_, s=1) |
| **ResBlock (C_in_, C_out_, s)** | Conv1d(C_in_, C_out_, k=3, s=s, p=1)  BatchNorm1d(C_out_)  ReLU  Conv1d(C_out_, C_out_, k=3, s=1, p=1)  BatchNorm1d(C_out_) |
| **TransformerLayer (d)** | Multi-Head Self-Attention(heads=8, d_k_=64, RoPE)  Dropout(0.1)  Residual connection  LayerNorm(d)  Linear(d, 4d) → GELU → Linear(4d, d)  Dropout(0.1)  Residual connection  LayerNorm(d) |

**Supplementary Results**

**Section 1. Additional Model Comparisons and Ablation Studies**

**1.1 Comparison with a silver-signal baseline**

To further assess whether the benefits of SleepFounder arise from its learned representations rather than simply from access to EEG- or oxygen-related information, we constructed an additional baseline. Specifically, SleepFounder was first used to generate silver EEG spectrograms and oxygen desaturation predictions from the original cardiorespiratory inputs (heartbeat and respiratory signals). These generated signals were then concatenated with the original cardiorespiratory inputs, yielding a four-channel input, and used to train a supervised model from scratch with an architecture identical to SleepFounder (except for the input layer). This baseline, denoted Silver Signals + Cardiorespiratory, tests whether the information captured during pretraining can be equivalently utilized through explicitly generated surrogate signals.

We evaluated both approaches across all four downstream tasks on both internal and external validation cohorts (Stanford, BIDMC, Emory, SSPH, SRRSH, and BCG). As shown in Supplementary Figure 2, SleepFounder consistently outperforms this silver-signal baseline on every individual dataset across all tasks, with average external results of Cohen’s kappa 0.649 vs. 0.624 for sleep staging, AUROC 0.915 vs. 0.902 for OSA detection, MAE 7.618 vs. 8.265 years for age prediction, and AUROC 0.888 vs. 0.850 for sex classification. The advantage holds uniformly across all internal and external cohorts, demonstrating that the pretraining stage confers benefits beyond what can be recovered by explicitly generating and appending silver-standard EEG and oxygen desaturation information.

We evaluated both approaches across four downstream tasks using internal and external cohorts (Stanford, BIDMC, Emory, SSPH, SRRSH, and BCG). As shown in Supplementary Figure 2, SleepFounder consistently outperformed the silver-signal baseline across all datasets and tasks, achieving average external performance of Cohen’s kappa 0.649 versus 0.624 for sleep staging, AUROC 0.915 versus 0.902 for OSA detection, MAE 7.618 versus 8.265 years for age prediction, and AUROC 0.888 versus 0.850 for sex classification. This performance advantage suggests that the benefits of pretraining extend beyond the information contained in silver-standard EEG and oxygen desaturation signals.

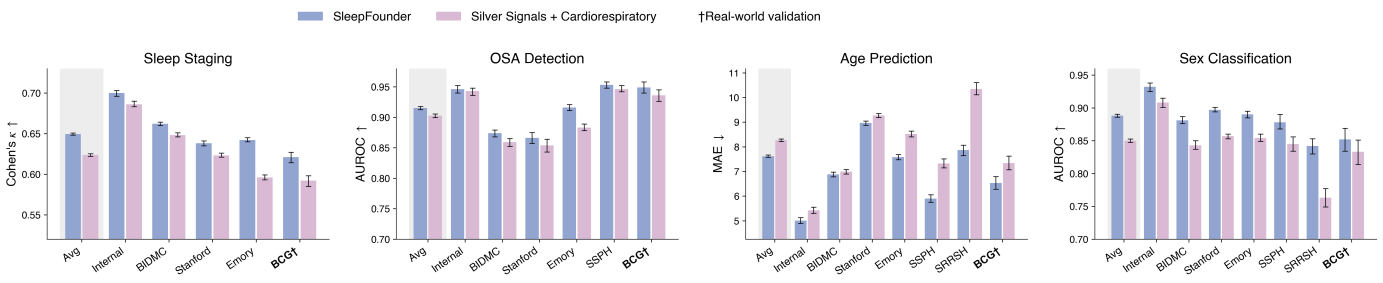
**Supplementary Figure 2. Comparison between SleepFounder and the Silver Signals + Cardiorespiratory baseline.** The baseline uses SleepFounder-generated silver EEG spectrograms and oxygen desaturation predictions concatenated with the original cardiorespiratory signals as inputs to a supervised model trained from scratch. Performance is reported across sleep staging, OSA detection, age prediction, and sex classification on both internal and external validation cohorts.

**1.2 Comparison with different time-series model architectures**

To evaluate the impact of model architecture, we compared SleepFounder against three representative time-series transformer backbones (FEDformer^9^, PatchTST^10^, and TimerXL^11^), using identical pretraining and fine-tuning protocols, training data, and evaluation splits. All models were trained on the same cardiorespiratory inputs and optimized under the same experimental settings.

As shown in Supplementary Figure 3, SleepFounder achieves competitive overall performance across the four downstream tasks. In terms of average metrics across all external sites, SleepFounder leads in sleep staging (Cohen’s kappa = 0.649 vs. FEDformer 0.642, PatchTST 0.637, TimerXL 0.626) and age prediction (MAE = 7.618 vs. FEDformer 7.818, PatchTST 8.193, TimerXL 8.473), while performing comparably to the best transformer baseline in OSA detection (AUROC 0.915 vs. FEDformer 0.919) and sex classification (AUROC 0.888 vs. FEDformer 0.890). Importantly, SleepFounder demonstrates substantially stronger generalization to the BCG device: on BCG cohorts, SleepFounder achieves sleep staging kappa = 0.621 (vs. next-best 0.610), OSA detection AUROC = 0.949 (tied with FEDformer, vs. PatchTST 0.929), age MAE = 6.533 (vs. next-best 8.025), and sex AUROC = 0.852 (vs. next-best 0.826). This cross-device robustness is particularly relevant given SleepFounder’s intended deployment on contactless devices, and demonstrates the advantage of our architecture over general-purpose time-series transformers for this application.

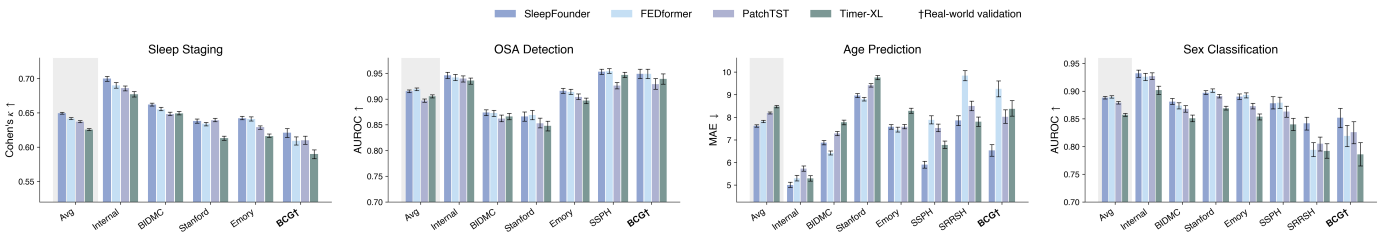
**Supplementary Figure 3. Comparison of SleepFounder with representative time-series transformer architectures (FEDformer, PatchTST, and TimerXL)**. All models were trained and evaluated using identical cardiorespiratory inputs, pretraining protocols, and downstream fine-tuning settings. Performance is reported across sleep staging, OSA detection, age prediction, and sex classification on both PSG and BCG cohorts.

1.3 **Comparison with supervised pretraining baseline method**

To evaluate the contribution of the proposed self-supervised pretraining strategy, we implemented a supervised pretraining baseline (SleepFounder-Supervised) using the same model architecture, training data, and optimization pipeline as SleepFounder. Specifically, the model was pretrained using joint supervision from all four downstream tasks (sleep staging, OSA detection, age prediction, and sex classification) before downstream fine-tuning.

As shown in Supplementary Figure 4, SleepFounder consistently outperformed SleepFounder-Supervised across all four tasks and validation cohorts. Based on average performance across external cohorts, SleepFounder achieved a Cohen’s kappa of 0.649 versus 0.624 for sleep staging, an AUROC of 0.915 versus 0.908 for OSA detection, an MAE of 7.618 versus 7.916 years for age prediction, and an AUROC of 0.888 versus 0.860 for sex classification. The performance advantage was consistently observed across both PSG and BCG evaluations, indicating that the proposed physiology-inspired self-supervised objectives learn representations that transfer more effectively across downstream tasks and sensing modalities than representations obtained through direct multi-task supervision.

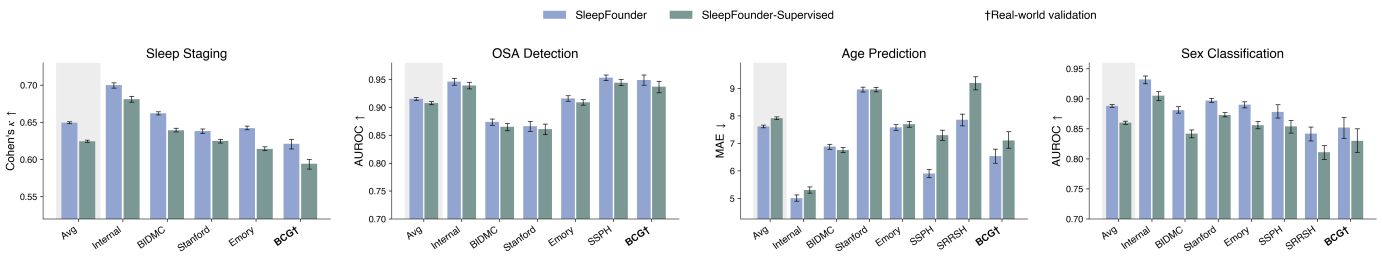
**Supplementary Figure 4. Comparison between SleepFounder and the SleepFounder-Supervised baseline.** The supervised baseline was pretrained using joint supervision from sleep staging, OSA detection, age prediction, and sex classification while using the same architecture, training data, and downstream evaluation protocol as SleepFounder. Performance is reported across PSG and BCG cohorts.

1.4 **Ablation of pretraining objectives**

To evaluate the contribution of each pretraining objective, we trained two single-task variants: SleepFounder-EEG, pretrained using EEG spectrogram reconstruction alone, and SleepFounder-SpO_2_, pretrained using oxygen desaturation prediction alone. Both variants were compared with the full dual-task SleepFounder under identical training and evaluation protocols.

As shown in Supplementary Figure 5, SleepFounder-EEG consistently achieved the weakest overall performance across downstream tasks, suggesting that EEG reconstruction alone provides a relatively limited pretraining signal. In contrast, SleepFounder-SpO_2_ achieved performance comparable to the full dual-task model on PSG-based external validation, with average performance of Cohen’s kappa = 0.646, AUROC = 0.916, MAE = 7.445 years, and AUROC = 0.889 across sleep staging, OSA detection, age prediction, and sex classification, compared with Cohen’s kappa = 0.649, AUROC = 0.915, MAE = 7.618 years, and AUROC = 0.888 for SleepFounder. The full model showed a modest advantage in sleep staging, consistent with the additional sleep-architecture information introduced through EEG reconstruction. Performance differences became more apparent during cross-device evaluation on contactless BCG recordings. SleepFounder achieved superior performance across all four downstream tasks, including sleep staging (Cohen’s kappa = 0.621 versus 0.608 for SleepFounder-SpO_2_ and 0.602 for SleepFounder-EEG), OSA detection (AUROC = 0.949 versus 0.945 and 0.929), age prediction (MAE = 6.533 years versus 6.671 and 8.214 years), and sex classification (AUROC = 0.852 versus 0.821 and 0.842). These findings suggest that the two pretraining objectives capture complementary physiological information and that combining both objectives yields representations that transfer more effectively across sensing modalities.

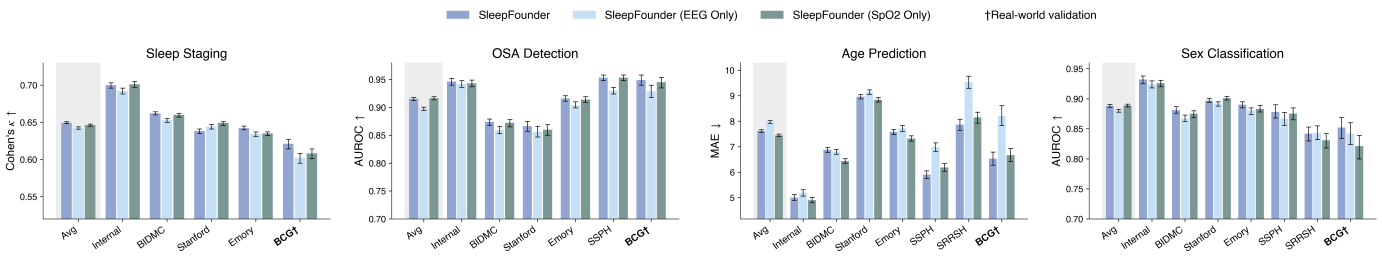
**Supplementary Figure 5. Ablation of pretraining objectives.** Comparison of SleepFounder with single-task variants pretrained using only oxygen desaturation prediction (SleepFounder-SpO_2_) or only EEG spectrogram reconstruction (SleepFounder-EEG). Performance is reported across sleep staging, OSA detection, age prediction, and sex classification on both PSG and BCG cohorts.

**1.5 Sensitivity analysis of loss weighting**

To assess the sensitivity of SleepFounder to the relative weighting of the two pretraining objectives, we compared the proposed loss weighting strategy with an equal-weight baseline (λ_EEG_ = λ_SpO2_) while keeping all other training settings unchanged.

As shown in Supplementary Figure 6, both weighting strategies achieved similar performance on PSG-based external validation. SleepFounder achieved average performance of Cohen’s kappa = 0.649, AUROC = 0.915, MAE = 7.618 years, and AUROC = 0.888 across the four downstream tasks, whereas the equal-weight variant achieved Cohen’s kappa = 0.658, AUROC = 0.913, MAE = 7.643 years, and AUROC = 0.890. A larger difference emerged during cross-device evaluation on BCG recordings. SleepFounder achieved Cohen’s kappa = 0.621, AUROC = 0.949, MAE = 6.533 years, and AUROC = 0.852, compared with Cohen’s kappa = 0.625, AUROC = 0.943, MAE = 7.321 years, and AUROC = 0.804 for the equal-weight variant. Although PSG performance was largely comparable, the proposed weighting strategy yielded more consistent performance under modality shift, particularly for demographic profiling tasks. Together with the pretraining-objective ablation results, these findings indicate that the selected loss weighting facilitates learning representations that generalize more effectively across sensing modalities.

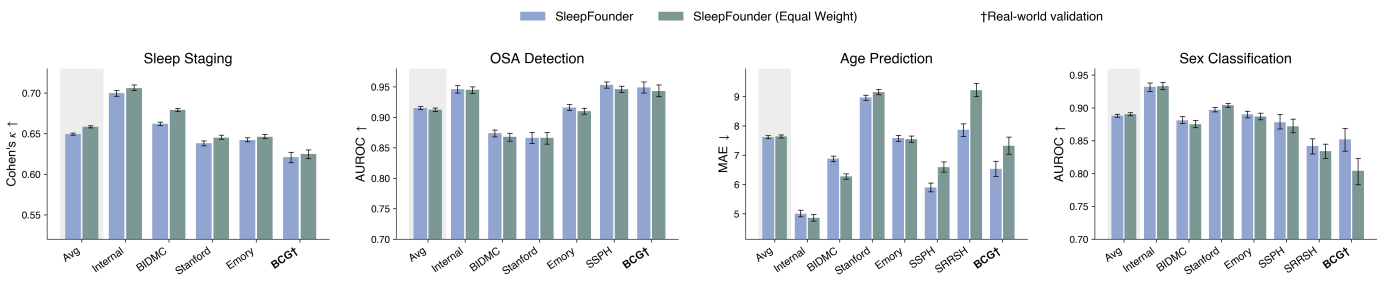
**Supplementary Figure 6. Sensitivity analysis of pretraining loss weighting**. Comparison between the proposed loss weighting strategy and an equal-weight baseline (λ_EEG_ = λ_SpO2_). Performance is reported across sleep staging, OSA detection, age prediction, and sex classification on both PSG and BCG cohorts.

**1.6 Comparison between SleepFM and SleepFM-CR**

As SleepFM-CR was adopted as a baseline in this study, we additionally compared the original SleepFM and SleepFM-CR using the same fine-tuning strategy described in the main text. External evaluation was performed on the BIDMC and Emory cohorts for sleep staging and age prediction. As shown in Supplementary Table 3, SleepFM-CR consistently outperformed the original SleepFM when using the publicly released checkpoints and model configurations from the original study. Therefore, SleepFM-CR was selected as the primary baseline, as it provides a more appropriate comparison for SleepFounder, which is designed to operate on cardiorespiratory signals for home-based and contactless sleep monitoring.

**Supplementary Table 3. External performance comparison between the original SleepFM and SleepFM-CR on sleep staging and age prediction tasks.** Both models were evaluated using the same fine-tuning strategy adopted in this study. Performance is reported using Cohen’s kappa for sleep staging and mean absolute error for age prediction. BIDMC, the Beth Israel Deaconess Medical Center

| Dataset | **Sleep Staging** | | **Age prediction** | |
| --- | --- | --- | --- | --- |
|  | SleepFM | SleepFM-CR | SleepFM | SleepFM-CR |
| BIDMC | 0.75 (0.74, 0.76) | 0.78 (0.77, 0.78) | 7.28 (6.97, 7.59) | 6.16 (5.9, 6.42) |
| Emory | 0.73 (0.72, 0.74) | 0.77 (0.76, 0.77) | 7.80 (7.48, 8.12) | 6.35 (6.09, 6.6) |

**Section 2. Additional Results for Phenome-Wide Disease Detection and Prediction**

**2.1 Disease prevalence and detailed per-site results**

Supplementary Data 2 provides, for each phecode phenotype included in the disease detection and future disease prediction analyses, the disease prevalence (or cumulative incidence for incident phenotypes), the number of positive cases, and detailed performance metrics for each external evaluation site (BIDMC, Stanford, and Emory).

In addition, we report the Area Under the Precision-Recall-Gain curve (AUPRG^12^) for all disease detection analyses (Supplementary Data 2). We selected AUPRG instead of the conventional area under the precision-recall curve (AUPRC) because AUPRG provides a prevalence-independent baseline of 0, enabling more meaningful comparison across phenotypes with substantially different prevalences. This property is particularly important in our phenome-wide evaluation, which spans 1,097 phenotypes covering a wide range of disease frequencies. As shown in Supplementary Figure 7, among the 172 phenotypes achieving an external average AUROC > 0.75, 158 (91.9%) also achieved AUPRG > 0, indicating performance above the no-skill baseline in the precision-recall domain. The median AUPRG among these high-performing phenotypes was 0.795 (IQR: 0.642-0.863), demonstrating that the observed discriminative performance is accompanied by meaningful precision-recall gains despite substantial class imbalance across diseases.

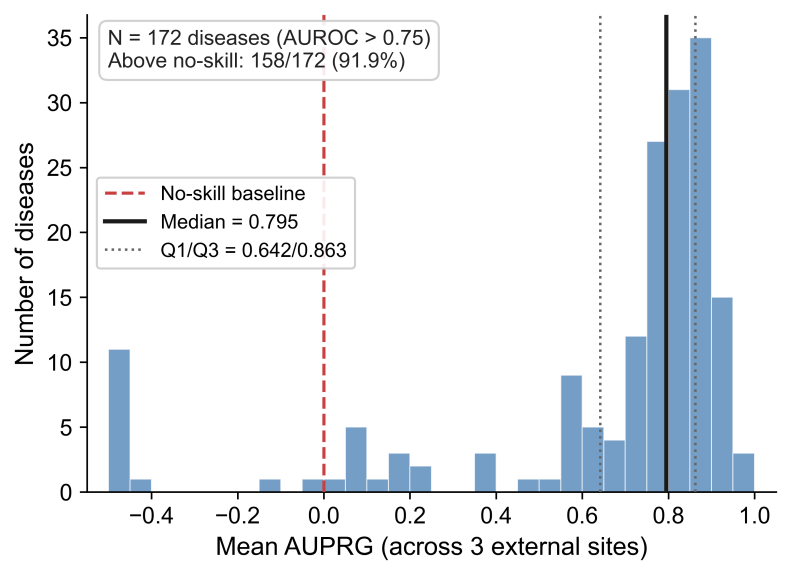

**Supplementary Figure 7. Distribution of AUPRG among high-performing disease phenotypes.** Histogram showing the distribution of external average Area Under the Precision-Recall-Gain curve (AUPRG) across the 172 disease phenotypes that achieved an external average AUROC > 0.75. Each bar represents the number of phenotypes within a given AUPRG interval. The red dashed line indicates the no-skill baseline (AUPRG = 0), the black solid line indicates the median AUPRG, and the gray dotted lines indicate the first and third quartiles (Q1/Q3). Among the 172 high-performing phenotypes, 158 (91.9%) achieved AUPRG values above the no-skill baseline, with a median AUPRG of 0.795 (IQR: 0.642-0.863), demonstrating that strong discriminative performance is generally accompanied by meaningful precision-recall gains despite substantial variation in disease prevalence.

**2.2 Phecode chapter-level disease detection and prediction results**

Supplementary Tables 4-5 summarize the chapter-level performance of SleepFounder for disease detection and future disease prediction, respectively. Consistent with the phenome-wide analyses presented in the main text, circulatory diseases exhibited the strongest performance, whereas phecode chapters with less direct expected coupling to overnight cardiorespiratory physiology, such as diseases of the sense organs and congenital anomalies, showed comparatively lower performance. Overall, the observed performance gradient across phecode chapters is broadly consistent with the known physiological relationships between sleep cardiorespiratory dynamics and multi-organ health.

Besides, Supplementary Tables 6-7 further summarize chapter-level disease detection and prediction performance for three models: SleepFounder with age and sex (as reported in the main text), SleepFounder without demographic variables, and a demographic baseline using age and sex only. Across most phecode chapters, the highest performance was achieved when SleepFounder representations were combined with age and sex, followed by SleepFounder alone, whereas the demographic baseline generally showed the lowest performance. Notably, SleepFounder without demographic information still achieved strong performance across a broad range of disease categories and substantially outperformed the demographic baseline in most phecode chapters. These findings indicate that the learned cardiorespiratory representations capture substantial disease-relevant information beyond demographic characteristics, while age and sex provide complementary information that further enhances predictive performance.

**Supplementary Table 4. Summary of disease detection performance of SleepFounder across phecode chapters (with age and sex as input).**

| **Chapter** | **N_diseases** | **Median AUROC** | **Q1** | **Q3** | **IQR** | **Min** | **Max** |
| --- | --- | --- | --- | --- | --- | --- | --- |
| VIII - Circulatory System | 134 | 0.730 | 0.689 | 0.784 | 0.689-0.784 | 0.494 | 0.855 |
| XI - Genitourinary | 43 | 0.691 | 0.638 | 0.737 | 0.638-0.737 | 0.507 | 0.851 |
| II - Endocrine/Metabolic | 102 | 0.687 | 0.627 | 0.762 | 0.627-0.762 | 0.552 | 0.854 |
| V - Mental Disorders | 67 | 0.669 | 0.634 | 0.738 | 0.634-0.738 | 0.536 | 0.884 |
| IV - Hematopoietic | 38 | 0.664 | 0.604 | 0.708 | 0.604-0.708 | 0.536 | 0.825 |
| XIII - Musculoskeletal | 84 | 0.652 | 0.604 | 0.701 | 0.604-0.701 | 0.447 | 0.825 |
| XV - Symptoms | 35 | 0.645 | 0.626 | 0.675 | 0.626-0.675 | 0.486 | 0.813 |
| XVI - Injuries & Poisonings | 65 | 0.644 | 0.604 | 0.695 | 0.604-0.695 | 0.474 | 0.829 |
| IX - Respiratory | 70 | 0.641 | 0.602 | 0.731 | 0.602-0.731 | 0.506 | 0.830 |
| I - Infectious Diseases | 43 | 0.640 | 0.602 | 0.705 | 0.602-0.705 | 0.507 | 0.796 |
| III - Neoplasms | 58 | 0.637 | 0.597 | 0.706 | 0.597-0.706 | 0.531 | 0.817 |
| VI - Neurological | 71 | 0.630 | 0.596 | 0.673 | 0.596-0.673 | 0.467 | 0.846 |
| X - Digestive | 122 | 0.627 | 0.595 | 0.650 | 0.595-0.650 | 0.458 | 0.784 |
| XII - Dermatologic | 69 | 0.625 | 0.580 | 0.692 | 0.580-0.692 | 0.542 | 0.837 |
| VII - Sense Organs | 75 | 0.617 | 0.566 | 0.674 | 0.566-0.674 | 0.444 | 0.826 |
| XIV - Congenital Anomalies | 21 | 0.616 | 0.590 | 0.665 | 0.590-0.665 | 0.470 | 0.785 |

**Supplementary Table 5. Summary of future disease prediction performance of SleepFounder across phecode chapters (with age and sex as input).**

| **Chapter** | **N_diseases** | **Median C-index** | **Q1** | **Q3** | **IQR** | **Min** | **Max** |
| --- | --- | --- | --- | --- | --- | --- | --- |
| VIII - Circulatory System | 134 | 0.713 | 0.669 | 0.747 | 0.669-0.747 | 0.547 | 0.816 |
| XI - Genitourinary | 43 | 0.670 | 0.613 | 0.724 | 0.613-0.724 | 0.434 | 0.816 |
| IV - Hematopoietic | 38 | 0.653 | 0.587 | 0.689 | 0.587-0.689 | 0.499 | 0.810 |
| II - Endocrine/Metabolic | 102 | 0.652 | 0.613 | 0.716 | 0.613-0.716 | 0.478 | 0.823 |
| V - Mental Disorders | 67 | 0.650 | 0.601 | 0.701 | 0.601-0.701 | 0.519 | 0.869 |
| XVI - Injuries & Poisonings | 65 | 0.644 | 0.596 | 0.677 | 0.596-0.677 | 0.467 | 0.810 |
| IX - Respiratory | 70 | 0.637 | 0.577 | 0.713 | 0.577-0.713 | 0.504 | 0.798 |
| III - Neoplasms | 58 | 0.636 | 0.585 | 0.677 | 0.585-0.677 | 0.503 | 0.788 |
| I - Infectious Diseases | 43 | 0.632 | 0.581 | 0.665 | 0.581-0.665 | 0.512 | 0.778 |
| XIII - Musculoskeletal | 84 | 0.619 | 0.587 | 0.661 | 0.587-0.661 | 0.514 | 0.780 |
| XV - Symptoms | 35 | 0.615 | 0.587 | 0.639 | 0.587-0.639 | 0.480 | 0.799 |
| VI - Neurological | 71 | 0.612 | 0.571 | 0.660 | 0.571-0.660 | 0.465 | 0.823 |
| X - Digestive | 122 | 0.608 | 0.585 | 0.636 | 0.585-0.636 | 0.455 | 0.742 |
| XII - Dermatologic | 69 | 0.607 | 0.555 | 0.653 | 0.555-0.653 | 0.520 | 0.820 |
| VII - Sense Organs | 75 | 0.591 | 0.551 | 0.637 | 0.551-0.637 | 0.453 | 0.814 |
| XIV - Congenital Anomalies | 21 | 0.586 | 0.562 | 0.619 | 0.562-0.619 | 0.497 | 0.649 |

**Supplementary Table 6. Chapter-level disease detection performance across model configurations. Results are summarized as median AUROC (Q1-Q3)。**AUROC, area under the receiver operating characteristic curve.

| **Chapter** | **N_diseases** | **SleepFounder + Age + Sex** | **SleepFounder** | **Age + Sex** |
| --- | --- | --- | --- | --- |
| VIII - Circulatory System | 134 | 0.730 (0.690-0.784) | 0.728 (0.684-0.777) | 0.668 (0.629-0.705) |
| XI - Genitourinary | 43 | 0.691 (0.638-0.737) | 0.664 (0.611-0.726) | 0.650 (0.569-0.680) |
| II - Endocrine/Metabolic | 102 | 0.687 (0.628-0.762) | 0.653 (0.589-0.750) | 0.624 (0.581-0.657) |
| V - Mental Disorders | 67 | 0.669 (0.634-0.738) | 0.659 (0.603-0.704) | 0.621 (0.580-0.682) |
| IV - Hematopoietic | 38 | 0.664 (0.604-0.708) | 0.654 (0.586-0.700) | 0.586 (0.564-0.633) |
| XIII - Musculoskeletal | 84 | 0.651 (0.604-0.701) | 0.619 (0.586-0.672) | 0.631 (0.587-0.705) |
| XV - Symptoms | 35 | 0.645 (0.625-0.675) | 0.625 (0.597-0.643) | 0.621 (0.587-0.637) |
| XVI - Injuries & Poisonings | 65 | 0.644 (0.604-0.695) | 0.627 (0.582-0.679) | 0.599 (0.557-0.635) |
| IX - Respiratory | 70 | 0.641 (0.602-0.731) | 0.642 (0.587-0.733) | 0.589 (0.562-0.632) |
| I - Infectious Diseases | 43 | 0.640 (0.603-0.706) | 0.619 (0.580-0.681) | 0.575 (0.531-0.609) |
| III - Neoplasms | 58 | 0.637 (0.597-0.706) | 0.625 (0.580-0.670) | 0.652 (0.604-0.709) |
| VI - Neurological | 71 | 0.630 (0.596-0.673) | 0.622 (0.585-0.664) | 0.581 (0.541-0.630) |
| X - Digestive | 122 | 0.627 (0.595-0.650) | 0.612 (0.581-0.634) | 0.588 (0.555-0.615) |
| XII - Dermatologic | 69 | 0.625 (0.580-0.691) | 0.603 (0.564-0.652) | 0.595 (0.565-0.645) |
| VII - Sense Organs | 75 | 0.617 (0.565-0.673) | 0.586 (0.548-0.624) | 0.603 (0.556-0.686) |
| XIV - Congenital Anomalies | 21 | 0.616 (0.590-0.665) | 0.594 (0.572-0.614) | 0.562 (0.500-0.651) |

**Supplementary Table 7. Chapter-level future disease prediction performance across model configurations. Results are summarized as median C-index (Q1-Q3)。**C-index, concordance index.

| **Chapter** | **N_diseases** | **SleepFounder + Age + Sex** | **SleepFounder** | **Age + Sex** |
| --- | --- | --- | --- | --- |
| VIII - Circulatory System | 134 | 0.713 (0.669-0.747) | 0.704 (0.662-0.739) | 0.658 (0.623-0.697) |
| XI - Genitourinary | 43 | 0.670 (0.613-0.724) | 0.650 (0.587-0.707) | 0.628 (0.577-0.657) |
| IV - Hematopoietic | 38 | 0.653 (0.587-0.689) | 0.645 (0.581-0.697) | 0.574 (0.548-0.622) |
| II - Endocrine/Metabolic | 102 | 0.652 (0.613-0.716) | 0.634 (0.569-0.699) | 0.613 (0.571-0.645) |
| V - Mental Disorders | 67 | 0.650 (0.600-0.701) | 0.620 (0.577-0.697) | 0.612 (0.571-0.677) |
| XVI - Injuries & Poisonings | 65 | 0.644 (0.596-0.677) | 0.631 (0.585-0.674) | 0.601 (0.560-0.627) |
| IX - Respiratory | 70 | 0.637 (0.577-0.713) | 0.638 (0.567-0.708) | 0.588 (0.554-0.643) |
| III - Neoplasms | 58 | 0.636 (0.586-0.677) | 0.613 (0.557-0.660) | 0.628 (0.573-0.678) |
| I - Infectious Diseases | 43 | 0.632 (0.581-0.665) | 0.600 (0.565-0.651) | 0.557 (0.516-0.609) |
| XIII - Musculoskeletal | 84 | 0.619 (0.587-0.660) | 0.595 (0.564-0.632) | 0.609 (0.579-0.645) |
| XV - Symptoms | 35 | 0.615 (0.588-0.639) | 0.591 (0.562-0.623) | 0.597 (0.569-0.612) |
| VI - Neurological | 71 | 0.612 (0.571-0.660) | 0.590 (0.573-0.640) | 0.571 (0.537-0.620) |
| X - Digestive | 122 | 0.608 (0.585-0.636) | 0.594 (0.574-0.625) | 0.572 (0.545-0.599) |
| XII - Dermatologic | 69 | 0.607 (0.555-0.653) | 0.586 (0.543-0.632) | 0.576 (0.534-0.631) |
| VII - Sense Organs | 75 | 0.591 (0.551-0.637) | 0.562 (0.544-0.611) | 0.588 (0.556-0.634) |
| XIV - Congenital Anomalies | 21 | 0.586 (0.562-0.619) | 0.564 (0.544-0.586) | 0.564 (0.539-0.595) |

**2.3 Disease detection performance in high-performing phecodes**

Supplementary Figure 8 summarizes disease detection performance for the 172 high-performing phecodes with an average external AUROC greater than 0.75. Performance was highly consistent between male and female participants, with a median AUROC difference of only 0.003. Larger differences were observed across age subgroups, with participants younger than 50 years achieving a median AUROC that was 0.032 higher than that of older participants; nevertheless, AUROC distributions remained consistently high in both age groups. Compared with a demographic baseline model using only age and sex as inputs, incorporating SleepFounder representations improved the median AUROC by 0.078 across the 172 phenotypes and achieved superior performance in 144 (83.7%) phecodes, highlighting the additional predictive value of the learned cardiorespiratory representations. Furthermore, performance within the OSA subgroup was largely consistent with that observed in the overall population, with a Spearman correlation coefficient of 0.718 and a median AUROC difference of only 0.009. Collectively, these findings indicate that SleepFounder maintains robust disease detection performance across demographic and clinical subgroups, suggesting that its predictive capability is not primarily driven by demographic characteristics or sleep-disordered breathing status alone.

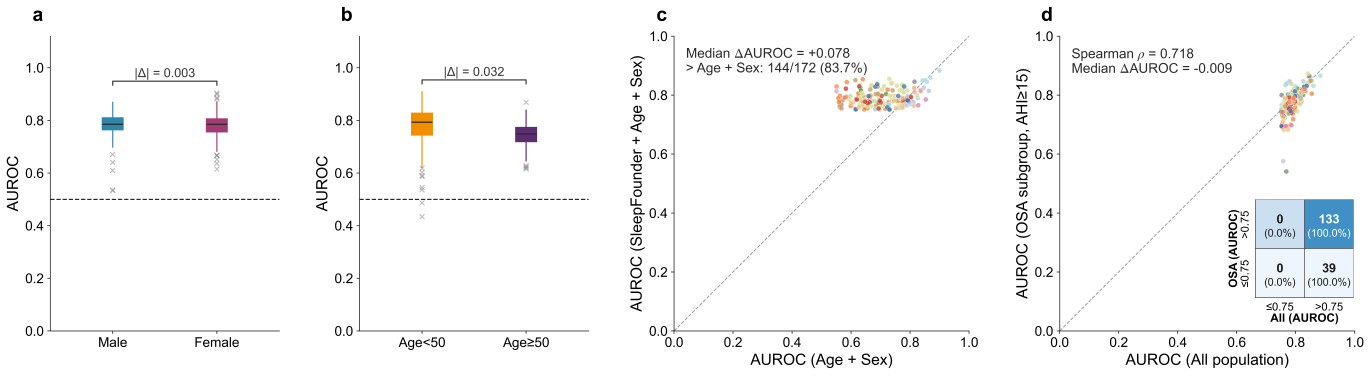

**Supplementary Figure 8. Disease detection performance among high-performing phenotypes (external average AUROC > 0.75).** (a) Comparison of external average AUROC distributions between male and female participants across the 172 high-performing phenotypes, showing highly consistent performance between sex subgroups, with a median AUROC difference of only 0.003. (b) Comparison of external average AUROC distributions between age subgroups (<50 years and ≥50 years), with the younger subgroup exhibiting a median AUROC 0.032 higher than the older subgroup, while maintaining consistently high performance in both groups. (c) Comparison of per-phenotype external average AUROC between SleepFounder (cardiorespiratory representations + age + sex) and a demographics-only baseline (age + sex). SleepFounder improved the median AUROC by 0.078 and achieved superior performance in 144 of 172 phenotypes (83.7%). (d) Per-phenotype comparison of external average AUROC between the overall population and the moderate-to-severe OSA subgroup analyses, showing strong agreement between the two settings (Spearman’s correlation coefficient = 0.718) and a median AUROC difference of only 0.009. AUROC, area under the receiver operating characteristic curve; OSA, obstructive sleep apnea.

**2.4 Representative calibration and decision curve analyses**

To further assess the reliability and potential clinical utility of the disease detection models, we performed calibration and decision curve analyses for six representative and clinically important diseases: congestive heart failure (AUROC = 0.840), atrial fibrillation (AUROC = 0.835), dementia (AUROC = 0.850), Parkinson’s disease (AUROC = 0.846), respiratory failure (AUROC = 0.822), and stage 4 chronic kidney disease (CKD; AUROC = 0.829). All analyses were based on pooled predictions from the three external validation cohorts (Stanford, BIDMC, and Emory), with site-specific results additionally examined to evaluate consistency across institutions. Calibration curves were constructed using 10 equal-frequency (quantile-based) bins. For all six diseases, the calibration curves closely approximated the ideal calibration line and exhibited a monotonic increase in observed event rates with increasing predicted risk. Expected calibration error (ECE) ranged from 0.0007 (stage 4 CKD) to 0.0257 (respiratory failure), whereas Brier scores ranged from 0.0145 to 0.0914 (Supplementary Figure 9), indicating generally good agreement between predicted and observed disease probabilities. Site-specific calibration analyses demonstrated similar patterns across the three external cohorts, further supporting the robustness of model calibration across institutions.

We further assessed potential clinical utility using decision curve analysis (Supplementary Figure 10), presenting prevalence-standardized net-benefit curves to facilitate comparisons across diseases with different baseline prevalences. Across the six representative diseases, the model generally provided greater net benefit than the “Treat All” and “Treat None” reference strategies over a broad range of low-to-intermediate decision thresholds. Similar trends were observed in the site-specific analyses, indicating that the potential clinical utility of the model was largely preserved across independent external cohorts.

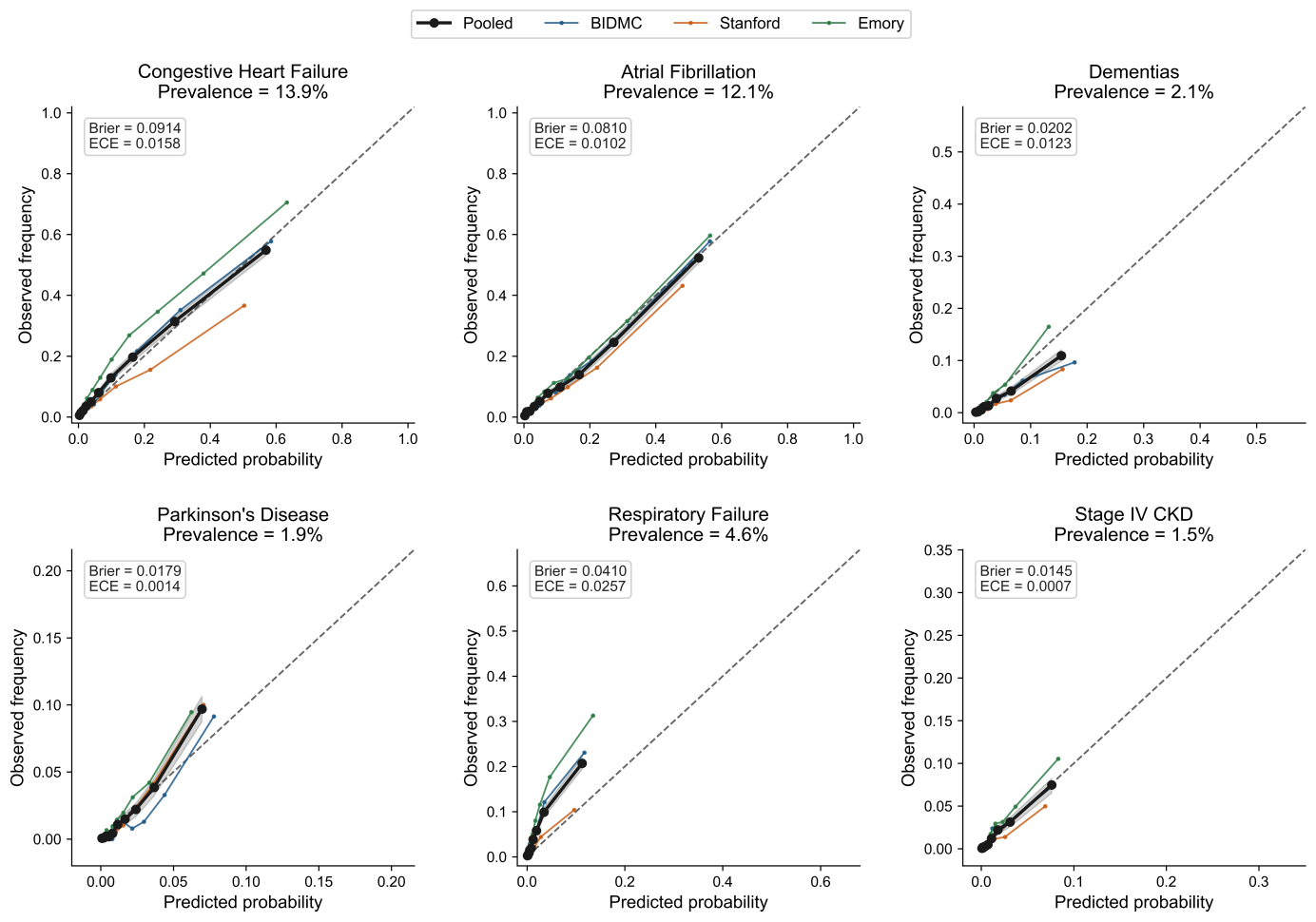

**Supplementary Figure 9. Calibration analysis for representative disease phenotypes.** Calibration curves for six representative diseases: congestive heart failure, atrial fibrillation, dementia, Parkinson’s disease, respiratory failure, and stage 4 CKD. Curves were generated using pooled predictions from the three external validation cohorts (Stanford, BIDMC, and Emory), with site-specific curves shown for comparison. The dashed diagonal line indicates perfect calibration. Expected calibration error (ECE) and Brier score are reported within each panel. BIDMC, the Beth Israel Deaconess Medical Center; CKD, chronic kidney disease.

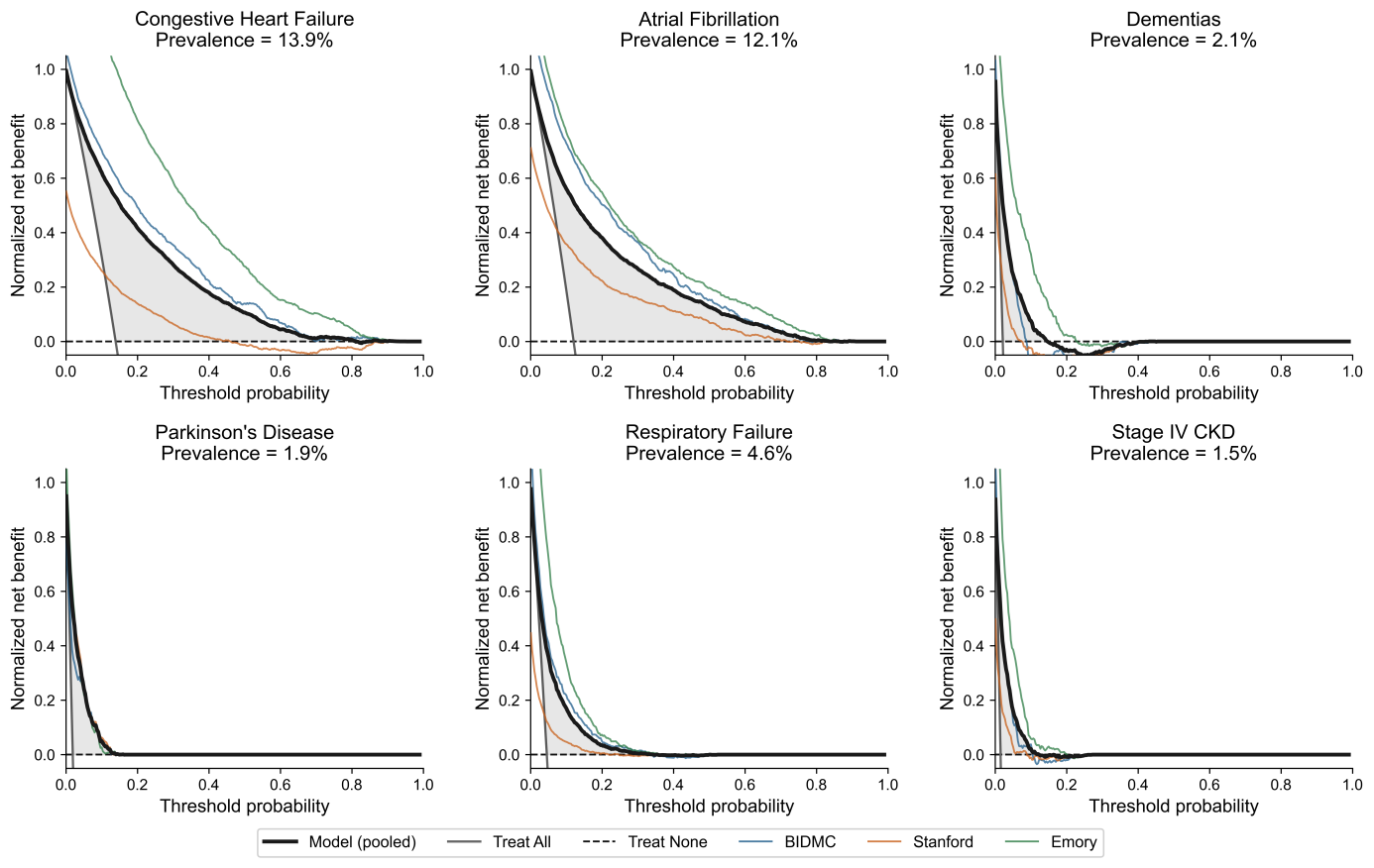

**Supplementary Figure 10. Decision curve analysis for representative disease phenotypes.** Prevalence-standardized net benefit curves for six representative diseases: congestive heart failure, atrial fibrillation, dementia, Parkinson’s disease, respiratory failure, and stage 4 CKD. Curves were generated using pooled predictions from the three external validation cohorts (Stanford, BIDMC, and Emory), with site-specific results shown for comparison. The “Treat All” and “Treat None” strategies are included as reference baselines. BIDMC, the Beth Israel Deaconess Medical Center; CKD, chronic kidney disease.

**2.5 Disease-specificity analysis using classifier-weight cosine similarity**

To further assess whether different disease phenotypes rely on distinct cardiorespiratory representation patterns, we computed pairwise cosine similarities between the classification-head weights of the 172 high-performing phenotypes (external average AUROC > 0.75). Results are summarized in Supplementary Figure 11. Overall, pairwise similarities were low, with a median cosine similarity of 0.135. No phenotype pair exhibited a cosine similarity greater than 0.5, and only 4.9% of phenotype pairs exceeded 0.25. Similarities between phenotypes within the same phecode chapter (median = 0.159) were modestly higher than the overall similarities across all phenotype pairs (median = 0.135), suggesting partially shared but largely distinct discriminative feature patterns among physiologically related diseases. Together, these findings support the disease-specific nature of the learned representations and indicate that SleepFounder does not rely on a single generic disease-discrimination signal across phenotypes.

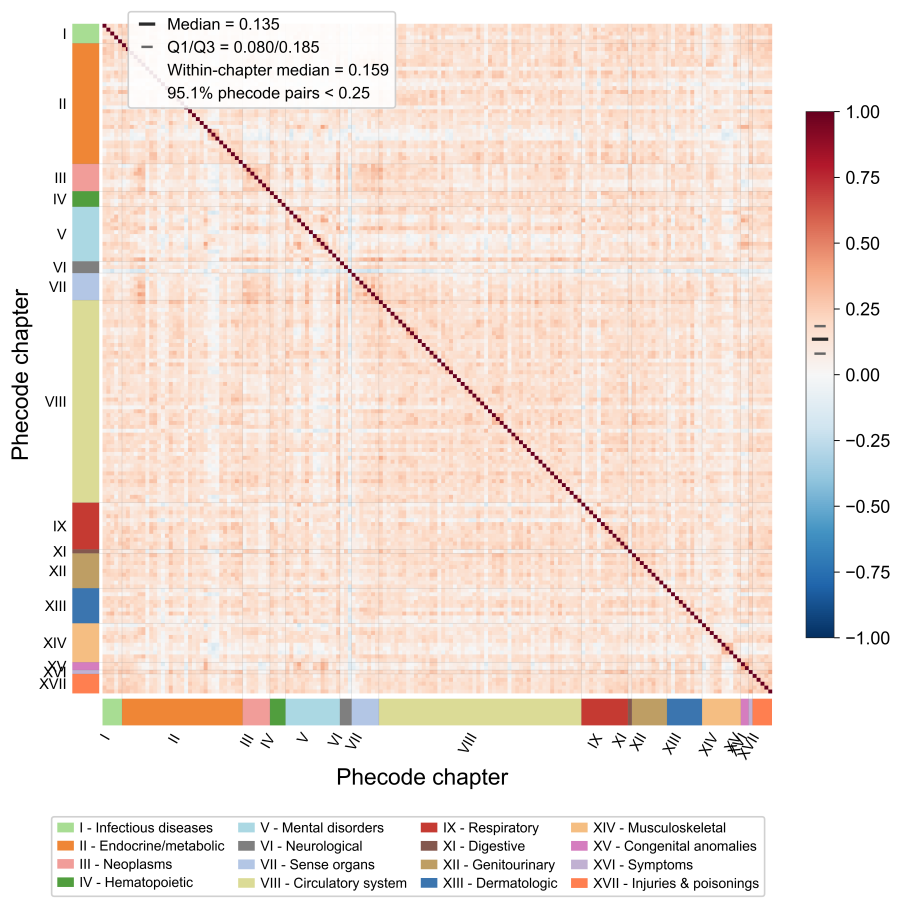

**Supplementary Figure 11. Pairwise cosine similarity among disease-specific classifier weight vectors.** Heatmap of pairwise cosine similarities computed from the normalized classifier weight vectors of the 172 high-performing disease phenotypes (external average AUROC > 0.75). Phenotypes are ordered by phecode chapter. The accompanying histogram summarizes the distribution of pairwise cosine similarities. Within-chapter similarity statistics are reported in the figure.

**2.6 Further Kaplan-Meier curves for future disease prediction**

Supplementary Figures 12-13 present additional Kaplan-Meier analyses from the BIDMC and Emory cohorts for representative phenotypes, including congestive heart failure, atrial fibrillation, respiratory failure, Parkinson’s disease, dementia, and stage 4 chronic kidney disease. Consistent with the findings in the Stanford cohort, the SleepFounder-predicted risk scores yielded clear separation of the event-free survival curves during follow-up, with significant differences between the high-risk and low-risk groups across all illustrated phenotypes (all log-rank P < 1e-10).

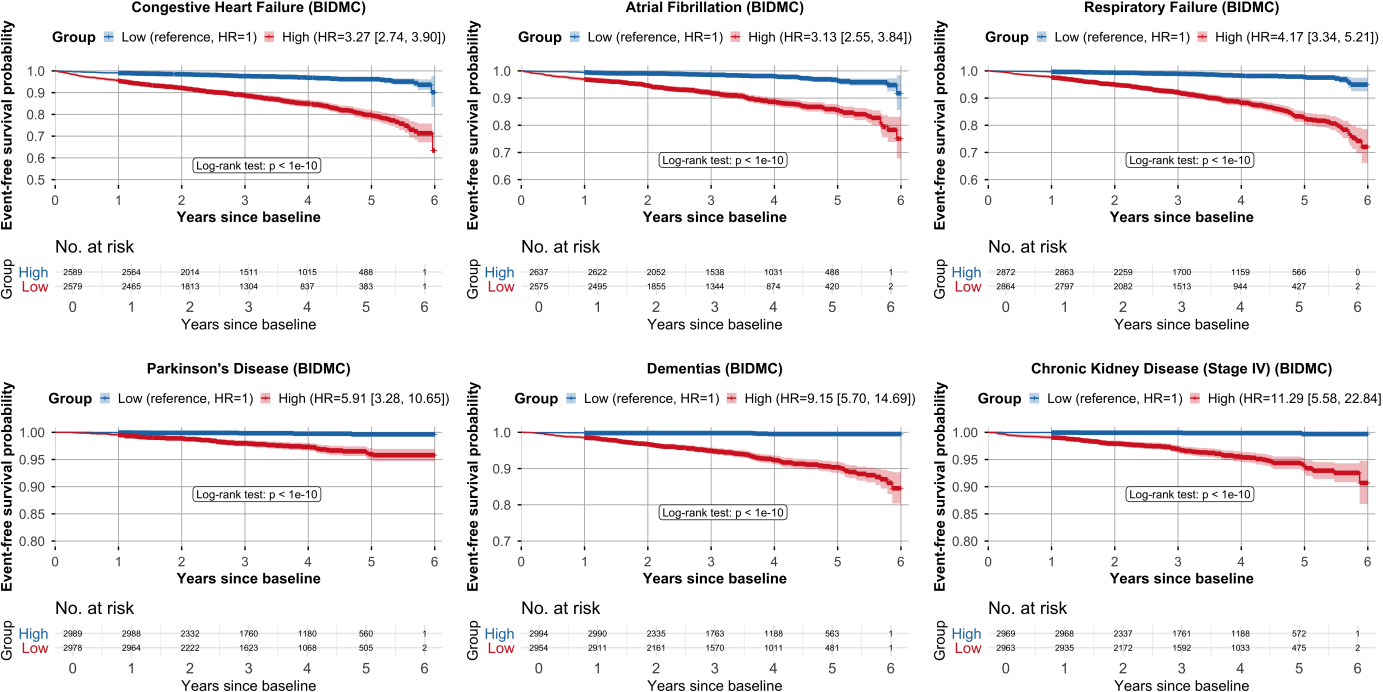

**Supplementary Figure 12. Kaplan-Meier analyses for representative phenotypes from the external BIDMC cohort, including congestive heart failure, atrial fibrillation, respiratory failure, Parkinson’s disease, dementia, and stage 4 chronic kidney disease.** Participants were stratified into high-risk and low-risk groups according to the median SleepFounder-predicted risk score (top 50% versus bottom 50% of the population). BIDMC, the Beth Israel Deaconess Medical Center; C-index, concordance index; HR, hazard ratio.

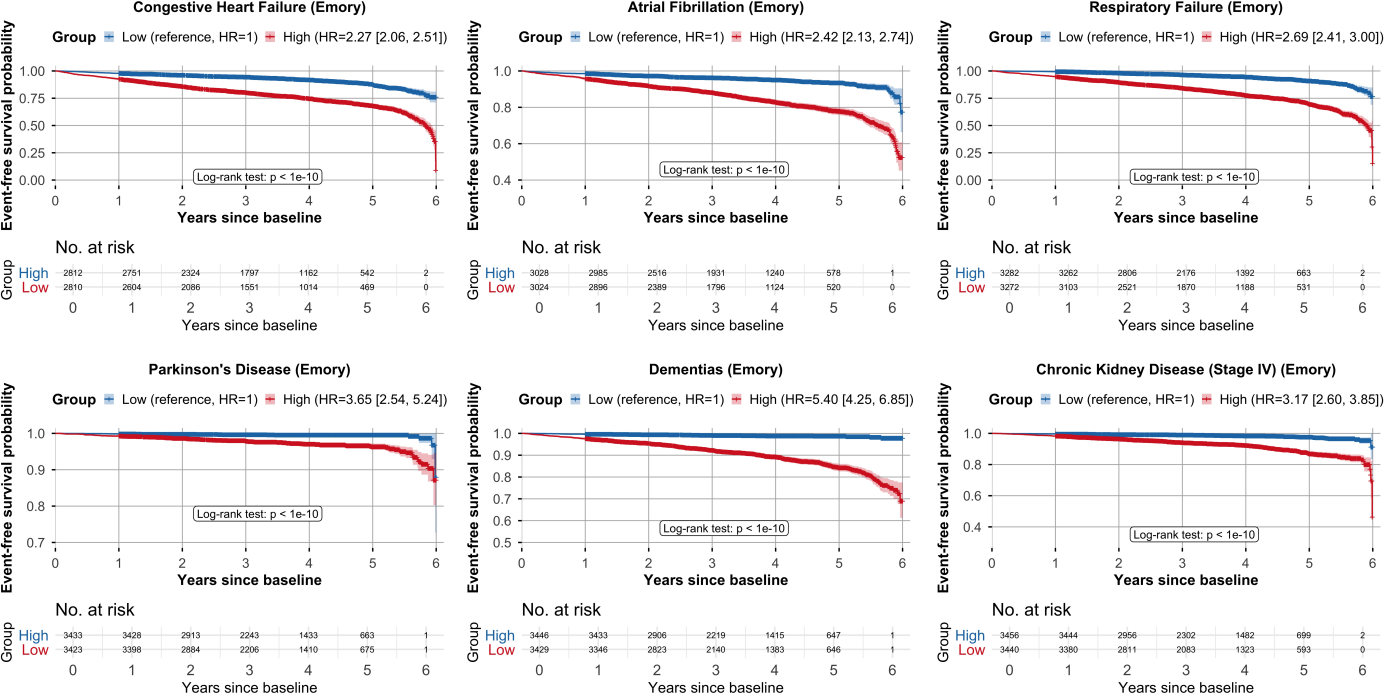

**Supplementary Figure 13. Kaplan-Meier analyses for representative phenotypes from the external Emory cohort, including congestive heart failure, atrial fibrillation, respiratory failure, Parkinson’s disease, dementia, and stage 4 chronic kidney disease.** Participants were stratified into high-risk and low-risk groups according to the median SleepFounder-predicted risk score (top 50% versus bottom 50% of the population). C-index, concordance index; HR, hazard ratio.

**Section 3. Comparison with Models Using Task-Specific PSG Channels**

To further assess the effectiveness of SleepFounder and the utility of cardiorespiratory signals, we compared its performance with existing task-specific models that employed full or task-relevant PSG channels across downstream tasks (Supplementary Table 8). Despite being trained exclusively on cardiorespiratory signals, SleepFounder achieved performance that was superior to, or only slightly below that of these task-specific models. This consistent performance across diverse sleep-related and disease prediction tasks highlights that cardiorespiratory dynamics alone capture sufficient physiological information for reliable and generalizable sleep and health assessment.

**Supplementary Table 8. Performance comparison between SleepFounder and existing approaches in sleep staging and age prediction.**

| Task | Method | Input Modality | Dataset | Performance (metric) |
| --- | --- | --- | --- | --- |
| Sleep staging | PFTSleep^13^ | PSG | SHHS | Kappa = 0.80 |
|  | XSleepNet2^14^ | PSG | SHHS | Kappa = 0.83 |
|  | SleepFM^5^ | PSG | SHHS | F1 = 0.78 |
|  | SleepFounder | Heartbeat, respiration | SHHS | Kappa = 0.74; F1 = 0.69 |
| Age prediction | Sun et al.^15^ | EEG | SHHS | SHHS1: MAE = 8.2 years  SHHS2: MAE = 9.4 years |
|  | Brink-Kjaer A et al.^16^ | ECG | SHHS | MAE = 10.4 years |
|  | **SleepFounder** | Heartbeat, respiration | SHHS | MAE = 5.1 years |

OSA, obstructive sleep apnea; PSG, polysomnography; EEG, electroencephalography; MAE, mean absolute error.
